# Cardiac dose-volume analysis of 9,411 patients with registry data for cardiovascular disease and overall survival

**DOI:** 10.1101/2024.08.16.24312108

**Authors:** Nora Forbes, Cynthia Terrones-Campos, Abraham George Smith, Joanne Reekie, Sune Darkner, Maja Maraldo, Mette Pøhl, Signe Risumlund, Lena Specht, Soren M. Bentzen, Jens Petersen, Ivan R. Vogelius

## Abstract

**Background and purpose:** Radiation therapy (RT) to the thorax poses risks of radiation-induced cardiotoxicity, potentially increasing cardiovascular diseases (CVD) incidence. Advances in RT strive to minimize these risks by reducing heart radiation dose exposure.

This study integrates detailed 3D dosimetry on individually delineated hearts with registry-based outcome data to assess the impact of radiation dose on cardiovascular morbidity and overall survival (OS) across multiple cancer types. It also examined the influence of patient-specific factors on cardiotoxicity risk and survival outcomes.

**Materials and methods:** We analyzed data from 9,411 patients receiving RT at Rigshospitalet between 2009 and 2020 for breast, esophageal, lymphoma, and lung cancers. Cumulative incidence of CVD and death in the presence of competing risks was calculated with the Aalen-Johansen estimator. The impact of radiation dose and patient characteristics on ischemic heart disease (IHD) onset and OS were assessed using Kaplan-Meier and Cox Proportional-Hazards Models.

**Results:** Higher mean heart dose (MHD) was associated with poorer OS in breast and lung cancer patients (Hazard ratio 2.8 and 1.2), but no significant relationship was found between MHD and IHD. Established cardiac risk factors (age, sex, and existing IHD) outweighed cardiac dose as a risk factor for subsequent cardiac events for all diagnoses. The risk of death was greater than subsequent CVD, especially in esophageal and lung cancers (cumulative incidence 60% versus 17% and 60% versus 14%), despite comparatively high heart doses.

**Conclusion:** The study demonstrates that risk of death from primary cancer is of far greater concern than risk of subsequent cardiac events from cardiac radiation dose exposure in the range achievable with contemporary RT techniques, especially for lung and esophageal cancer patients. Further sparing of the heart should not be prioritized at the expense of adequate treatment of the index cancer.

**Highlights:**

- Age and existing heart disease far outweighed heart dose as predictors of ischemic heart disease
- Overall survival is not a useful surrogate for cardiac toxicity in dose-response studies due to confounding by disease stage
- With modern RT techniques, the excess absolute risk attributable to radiotherapy is so small that a statistically significant dose-response could not be observed even in 9,411 patients
- For most patients, good quality contemporary radiotherapy is sufficient to limit heart toxicity as a clinically relevant concern

## Introduction

Radiation therapy (RT) is a critical treatment modality for cancer, offering effective control for many tumor types. Despite therapeutic benefits, RT can pose significant risks of adverse events from unavoidable irradiation of normal tissue surrounding the target. Radiation-induced cardiotoxicity is a concern among thoracically irradiated patients [1,2]. It can manifest as ischemic heart disease (IHD) among other cardiovascular diseases (CVD).

Radiation-induced cardiotoxic late-effects are an increasing concern for cancer patients [3–10]. The focus on cardiotoxicity mitigation is increasing with the establishment of guidelines including thresholds on delivered cardiac dose [11]. Darby et. al quantified the risk of CVD in breast cancer patients in 2013; recent work has extended findings into lymphoma and lung cancers [10,12]. Ongoing advances in radiation technology and techniques aim to reduce CVD risks by minimizing the mean heart dose (MHD) and other critical dosimetric parameters. These measures resulted in significant decreases in cardiac dose [13].

Many studies highlight the complex interplay between radiation dose, patient characteristics, and cancer stage on cardiac outcomes [14]. Particularly, the relationship between radiation-induced cardiotoxicity and overall survival (OS) has garnered attention, with evidence suggesting higher radiation doses to the heart correlate with poorer survival outcomes in certain cancers [15–18]. Therefore, there is a need for precise dosimetric assessments and comprehensive outcome data to understand the dose effect on overt cardiotoxicity.

The current study integrates detailed 3D dosimetry data on 9,411 individually delineated hearts with outcomes from national registry systems to analyze cardiotoxicity and mortality following standard cancer treatment with RT. We aimed to quantify and contextualize the impact of radiation dose and patient-specific factors on cardiotoxic side effects and OS. These findings can inform optimal directions for future refinement of treatment planning and delivery.

## Methods

### Data Curation

Eligible patients included those treated with curative intent at Rigshospitalet in 2009-2020. Patients with breast cancer, esophageal cancer, lymphoma, and non-small cell (NSCLC) or small cell (SCLC) lung cancer were included. We identified patients by departmental codes used in the record and verify system.

Associated computed tomography (CT) scans and 3D dose matrices were acquired through the record and verify system. Heart segmentation on all planning CT scans were performed using an open-source AI software [19]. All heart delineations were validated by manual review in 2D. Questionable cases were secondarily reviewed in 3D with a physician and unacceptable cases were omitted. Calculated dose metrics included mean heart dose (MHD), absolute volume receiving at least 5 Gy (V5), and absolute volume receiving at least 30 Gy (V30) - all converted to equivalent doses in 2 Gy (EQD2) fractions (α/β ratio = 2 Gy). Sex and birthdate were collected from patient records, as previously detailed in Forbes et al. 2024 [13].

Outcome data were extracted through electronic health records (EHR) maintained at the Centre of Excellence for Personalized Medicine of Infectious Complications in Immune Deficiency (PERSIMUNE) data warehouse. Diagnostic codes were sourced from The National Patient Register (Landspatientregisteret (LPR)) database. Death and emigration records were sourced from the Central Person Register (CPR).

### Statistical Analysis

A statistical analysis plan (SAP) was established and published prior to analysis [20]. Modifications were made to the original SAP (Table A1).

The primary endpoint was onset of IHD following RT, defined by ICD-10 codes I20-I25 occurring after baseline, defined as the date of the last fraction of RT. Additionally, a broader outcome of CVD including valvular disease (VD; I00-I09, I34-I39) and heart failure (HF; I50) was assessed. OS was also analyzed.

All analyses were conducted separately on each cancer diagnosis due to heterogeneity among patient populations. The reverse Kaplan-Meier (KM) method was used to report follow-up in the absence of an event [21]. Absolute risk estimates of the competing events of the three CVD outcomes and death from other causes were analyzed using Aalen-Johansen estimates.

To predict risk of IHD, KM and Cox Proportional-Hazards Model (PHM) were used. Univariable KM curves were generated for each independent variable. Survival distributions were compared between factor levels with a log rank test (for variables with 2 levels) or test for trend analysis (for variables with >2 levels). Multivariable analysis of these variables was conducted with the Cox PHM. Patients were censored at the date of emigration, death, or the last potential follow-up (December 31^st^, 2020). These KM and Cox PHM were then replicated with OS as the outcome.

Independent variables of interest included sex, age, existing IHD, and MHD. Sex was omitted for the breast cancer cohort as the very few males were excluded. Age was calculated at baseline and fit as a categorical variable with three levels (<60, 60-70, and >70). Existing IHD was defined as the same diagnostic codes as the outcome occurring prior to baseline. For the KM analyses, MHD was presented as below or above the group specific median for visual simplicity. For the Cox PHM, MHD was fitted as a linear predictor.

Sub-analyses were conducted on populations with and without existing IHD as a secondary analysis and among lymphoma patients receiving high MHD (>5 Gy). Young patients in the breast and lymphoma cohorts were further subdivided into age groups (<40, 40-50, and 50-60), due to their abundance and heterogeneity. The KM and Cox PHM were replicated with the broader definition of diagnosis codes to represent CVD in place of IHD. This was for both the outcome and independent variable of pre-existing disease. The impact of including a low (V5) and high (V30) volumetric dose measure on model performance was assessed with log-rank tests comparing nested Cox PHMs.

All analyses were conducted using R statistical software [22,23]. Packages ‘prodlim’, ‘survival’, and ‘rms’ were used to conduct analysis [24–26]. Additionally, we leveraged ‘ggplot2’, ‘ggfortify’, ‘sjPlot’, and ‘cowplot’ to generate figures [27–30]. p<0.05 was considered significant.

## Results

9,411 patients were included in the outcome analysis (Figure 1). Prior cancer was the main reason for exclusion. Breast cancer formed the largest proportion with 6808 patients, followed by lymphoma with 1251, esophageal with 627, and lung with 729. Breast cancer and lymphoma had the youngest populations, see Table 1. In terms of dosimetric data, MHD was less than 1.74 Gy in 90% of breast cancer cases.

**Figure 1.**
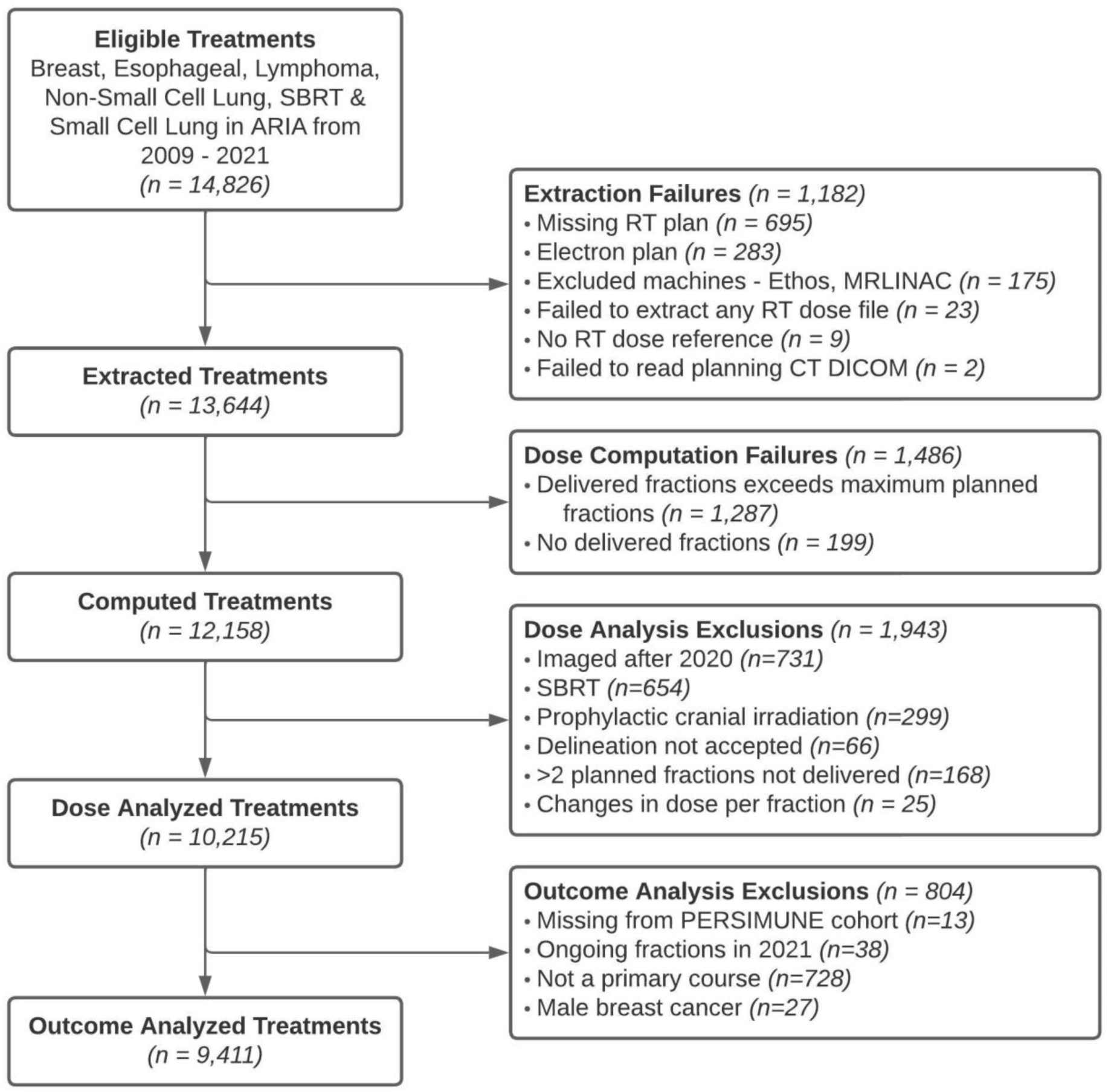
Consort diagram.

**Table 1.**
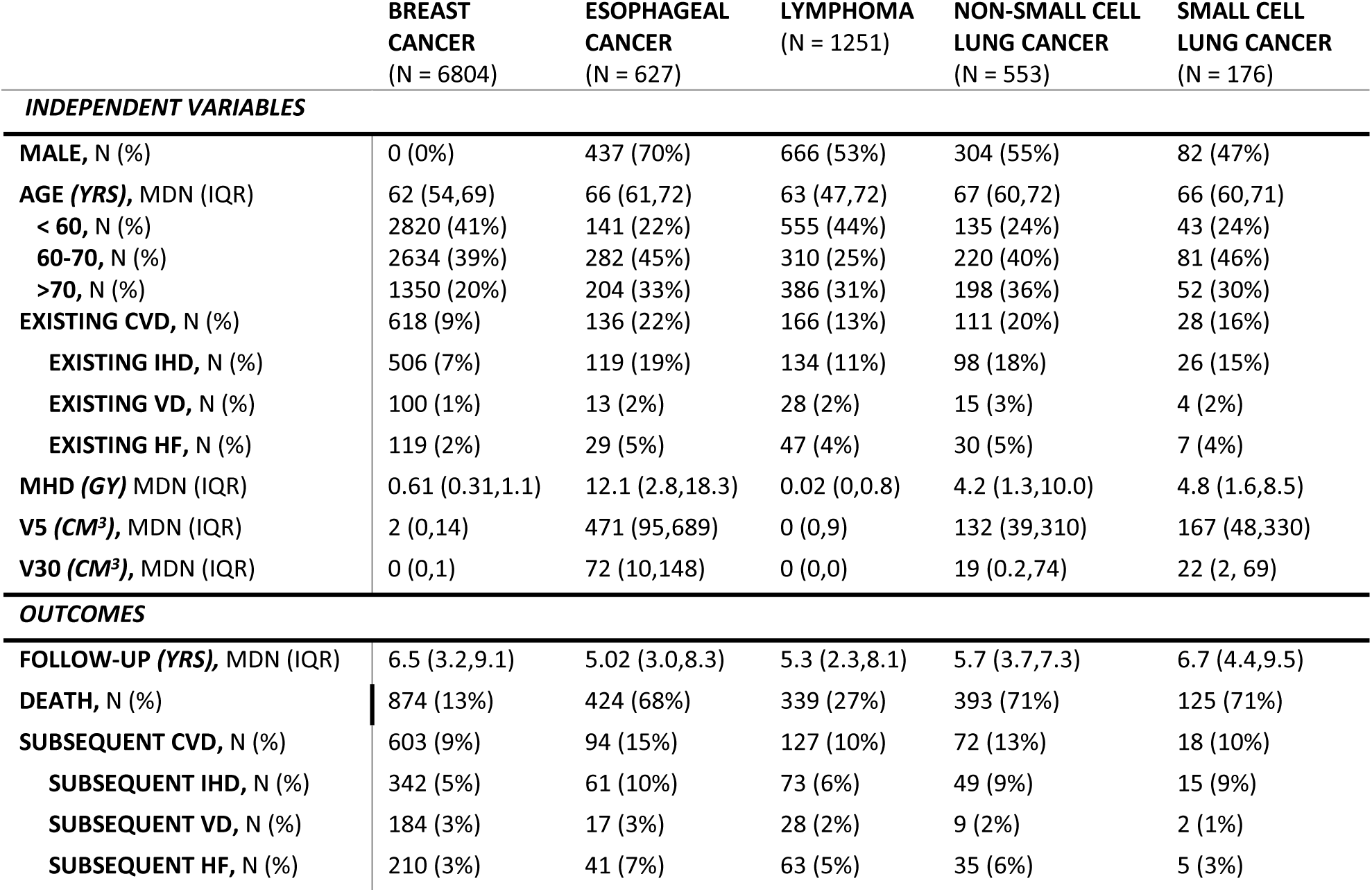
Patient characteristics presented as independent variables – sex, age, existing cardiovascular disease (CVD), and dose metrics – and outcomes – subsequent CVD, death, and follow-up time. CVD includes ischemic heart disease (IHD), valvular disease (VD), and heart failure (HF). Existing CVD is defined as occurring before baseline (delivery date of last fraction) and subsequent CVD is defined as occurring after baseline. Dose metrics include mean heart dose (MHD), absolute volume receiving at least 5 Gy (V5), and absolute volume receiving at least 30 Gy (V30). Doses are presented in equivalent dose in 2 Gy fractions (EQD2). Continuous variables are presented as the median (MDN) and interquartile range (IQR). Categorical variables are presented as the number (N) and percentage (%) of patients. Follow-up time to first event is calculated using the reverse Kaplan-Meier estimator.

|  | **BREAST**<br>**CANCER**<br>**(N = 6804)** | **ESOPHAGEAL**<br>**CANCER**<br>**(N = 627)** | **LYMPHOMA**<br>**(N = 1251)** | **NON-SMALL CELL**<br>**LUNG CANCER**<br>**(N = 553)** | **SMALL CELL**<br>**LUNG CANCER**<br>**(N = 176)** |
| --- | --- | --- | --- | --- | --- |
| **INDEPENDENT VARIABLES** |  |  |  |  |  |
| **MALE, N (%)** | 0 (0%) | 437 (70%) | 666 (53%) | 304 (55%) | 82 (47%) |
| **AGE (YRS), MDN (IQR)** | 62 (54,69) | 66 (61,72) | 63 (47,72) | 67 (60,72) | 66 (60,71) |
| **< 60, N (%)** | 2820 (41%) | 141 (22%) | 555 (44%) | 135 (24%) | 43 (24%) |
| **60-70, N (%)** | 2634 (39%) | 282 (45%) | 310 (25%) | 220 (40%) | 81 (46%) |
| **>70, N (%)** | 1350 (20%) | 204 (33%) | 386 (31%) | 198 (36%) | 52 (30%) |
| **EXISTING CVD, N (%)** | 618 (9%) | 136 (22%) | 166 (13%) | 111 (20%) | 28 (16%) |
| **EXISTING IHD, N (%)** | 506 (7%) | 119 (19%) | 134 (11%) | 98 (18%) | 26 (15%) |
| **EXISTING VD, N (%)** | 100 (1%) | 13 (2%) | 28 (2%) | 15 (3%) | 4 (2%) |
| **EXISTING HF, N (%)** | 119 (2%) | 29 (5%) | 47 (4%) | 30 (5%) | 7 (4%) |
| **MHD (GY) MDN (IQR)** | 0.61 (0.31,1.1) | 12.1 (2.8,18.3) | 0.02 (0,0.8) | 4.2 (1.3,10.0) | 4.8 (1.6,8.5) |
| **V5 (CM<sup>3</sup>), MDN (IQR)** | 2 (0,14) | 471 (95,689) | 0 (0,9) | 132 (39,310) | 167 (48,330) |
| **V30 (CM<sup>3</sup>), MDN (IQR)** | 0 (0,1) | 72 (10,148) | 0 (0,0) | 19 (0.2,74) | 22 (2, 69) |
| **OUTCOMES** |  |  |  |  |  |
| **FOLLOW-UP (YRS), MDN (IQR)** | 6.5 (3.2,9.1) | 5.02 (3.0,8.3) | 5.3 (2.3,8.1) | 5.7 (3.7,7.3) | 6.7 (4.4,9.5) |
| **DEATH, N (%)** | 874 (13%) | 424 (68%) | 339 (27%) | 393 (71%) | 125 (71%) |
| **SUBSEQUENT CVD, N (%)** | 603 (9%) | 94 (15%) | 127 (10%) | 72 (13%) | 18 (10%) |
| **SUBSEQUENT IHD, N (%)** | 342 (5%) | 61 (10%) | 73 (6%) | 49 (9%) | 15 (9%) |
| **SUBSEQUENT VD, N (%)** | 184 (3%) | 17 (3%) | 28 (2%) | 9 (2%) | 2 (1%) |
| **SUBSEQUENT HF, N (%)** | 210 (3%) | 41 (7%) | 63 (5%) | 35 (6%) | 5 (3%) |

The median follow-up time was 6.3 years (IQR 3.1-8.9). The 3-year cumulative incidence [95% CI] of CVD was greatest at 17% [13-21%] in esophageal, followed by 14% [11-17%] in lung, 9% [7-11%] in lymphoma, and 5.3% [5-6%] breast cancer patients (Figure 2). The 3-year cumulative incidence of death far outweighed that of CVD, especially in esophageal at 60% [56-64%] and in lung at 60% [56-63%], and to a lesser extent at 20% [17-22%] in lymphoma and 5% [4-5%] in breast cancer patients. Patients with breast cancer had a steady linearly increasing risk of death across time, while the other patient groups had a higher initial risk of death in the first year.

**Figure 2.**
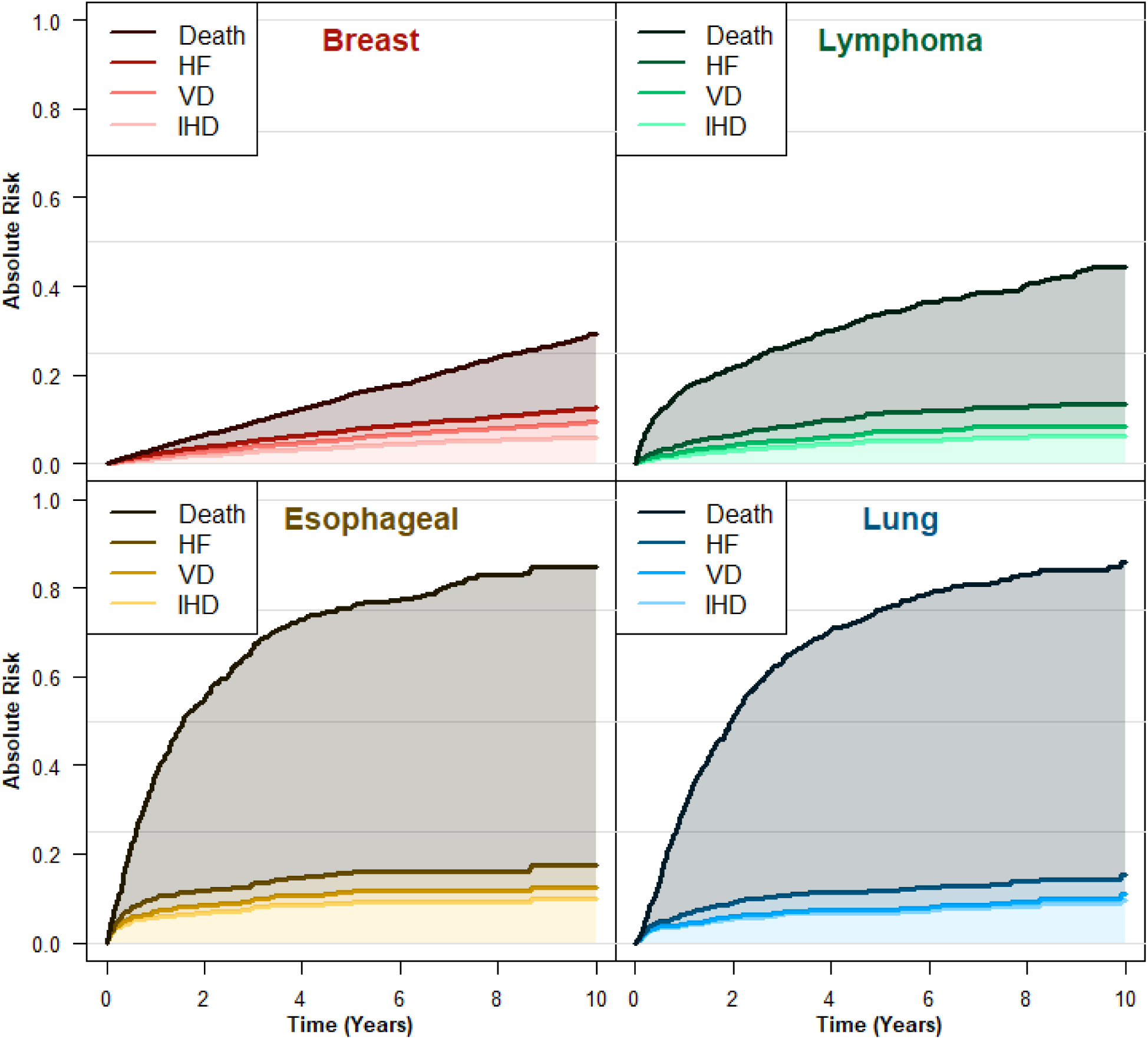
Stacked cumulative incidence plots for each outcome – death, heart failure (HF), valvular disease (VD), and ischemic heart disease (IHD), separated by diagnosis. Numbers at risk available in supplementary Tables A4–6.

Existing IHD had a large impact on subsequent IHD across all groups (Figure 3) and was associated with poor OS in all but lung cancer patients (Figure 4). In contrast, the effect of MHD on IHD was small and non-significant, except for patients with lymphoma where the relationship was inverse, see Figure 3. A high MHD was significantly associated with poorer OS in breast and lung cancer patients, see Figure 4. Patients with lymphoma and a MHD above 5 Gy were also significantly younger than those with a lower MHD (median age of 44.9 compared to 63.3; P < 0.001 (Table A2)). Age had a significant impact on IHD in patients with breast cancer and lymphoma, and on OS in all cancers except esophageal.

**Figure 3.**
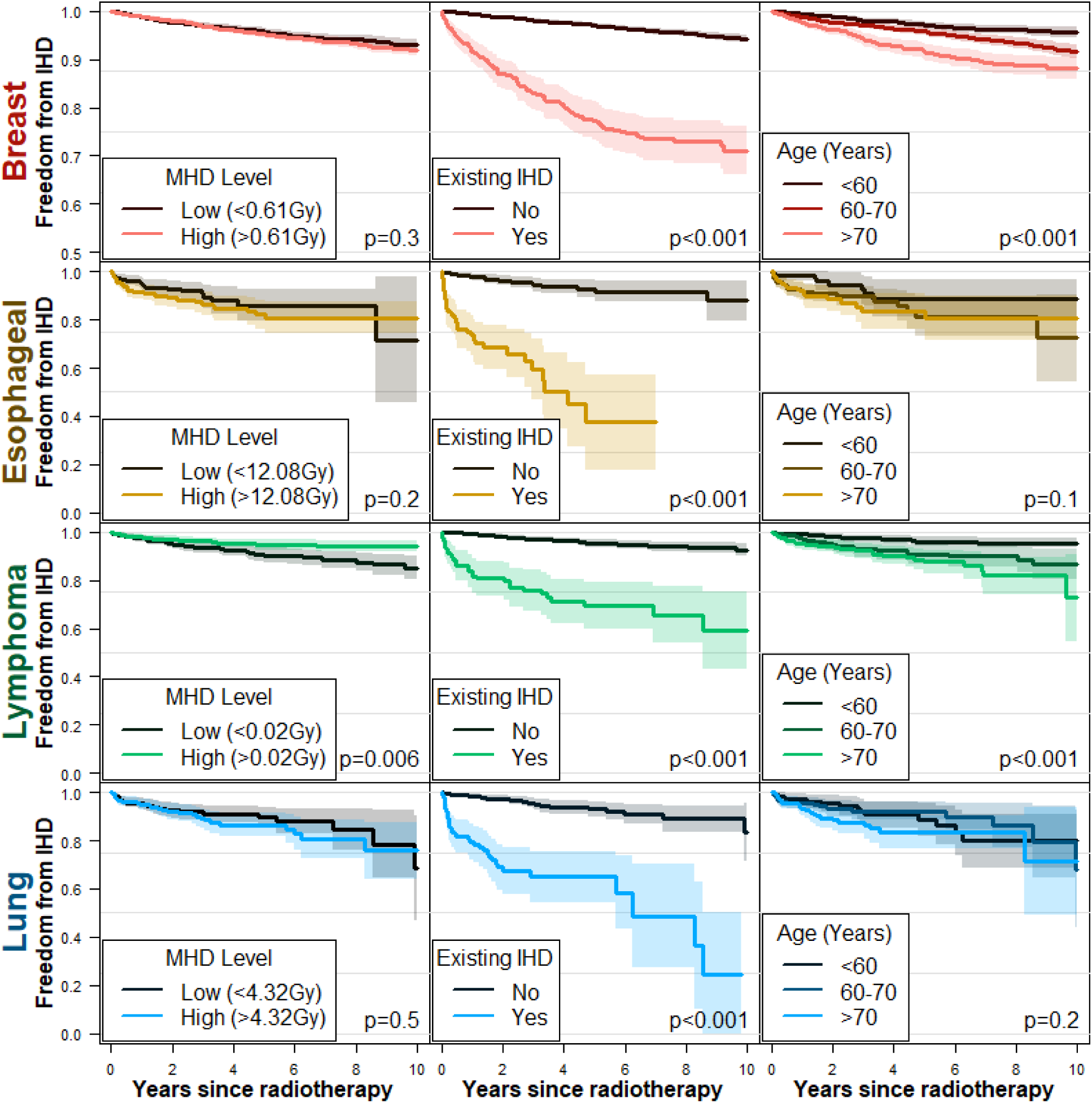
Kaplan-Meier plots for freedom from ischemic heart disease (IHD), by mean heart dose (MHD) level (below or above median), existing IHD, and age group (<60, 60-70, and >70 years old), separated by diagnosis. P-values generated from log-rank test (MHD level and existing IHD) and test for trend (age group). *Note that the Breast cancer y-axis is truncated at 0.5 to better show separation.* Numbers at risk available in supplementary Table A4. KM plots for overall CVD available in supplement Figure A7. For the effect of sex on freedom from IHD see supplementary Figure A1.

**Figure 4.**
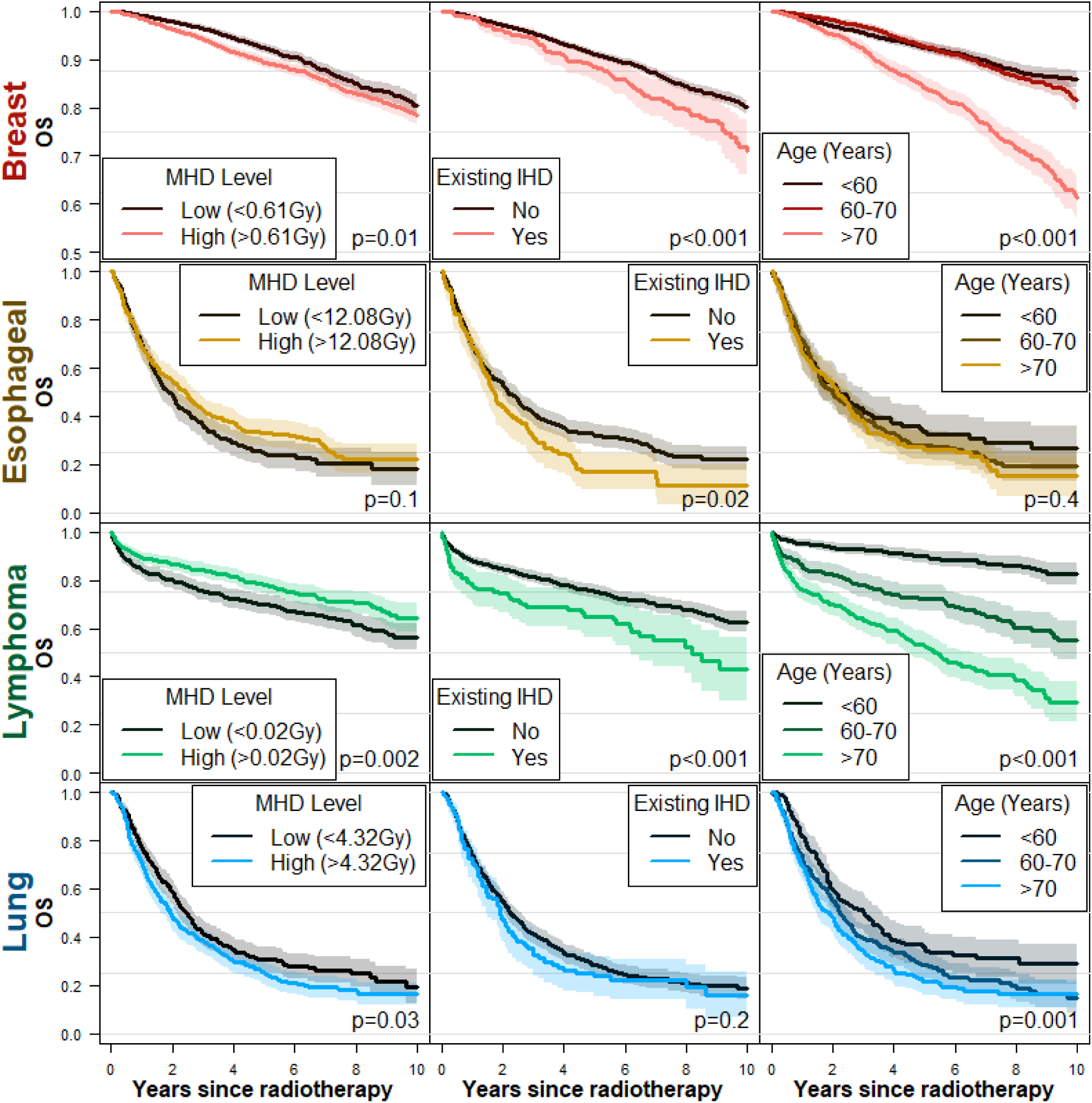
Kaplan-Meier plots for overall survival (OS), by mean heart dose (MHD) level (below or above median), existing ischemic heart disease (IHD), and age group (<60, 60-70, and >70 years old), separated by diagnosis. *Note that the Breast cancer y-axis is truncated at 0.5 to better show separation*. P-values generated from log-rank test (MHD level and existing IHD) and test for trend (age group). Numbers at risk available in supplement Table A5. For the effect of sex on OS see supplementary Figure A1.

The multivariable analysis demonstrated how the magnitude of existing IHD far outweighed the other predictors for subsequent IHD, see Figure 5. MHD was not significantly associated with IHD in any group. There was a large significant effect of MHD on OS in patients with breast cancer (HR 2.8) and a smaller, but still significant effect of high MHD on OS in patients with lung cancer (HR 1.2). Age was significantly associated with OS for all groups apart from esophageal cancer, while the effect was particularly strong in lymphoma patients. There was no significant effect of sex in any group for either outcome, though the direction of the effect was as expected with males having higher hazards than females (Figure A1).

**Figure 5.**
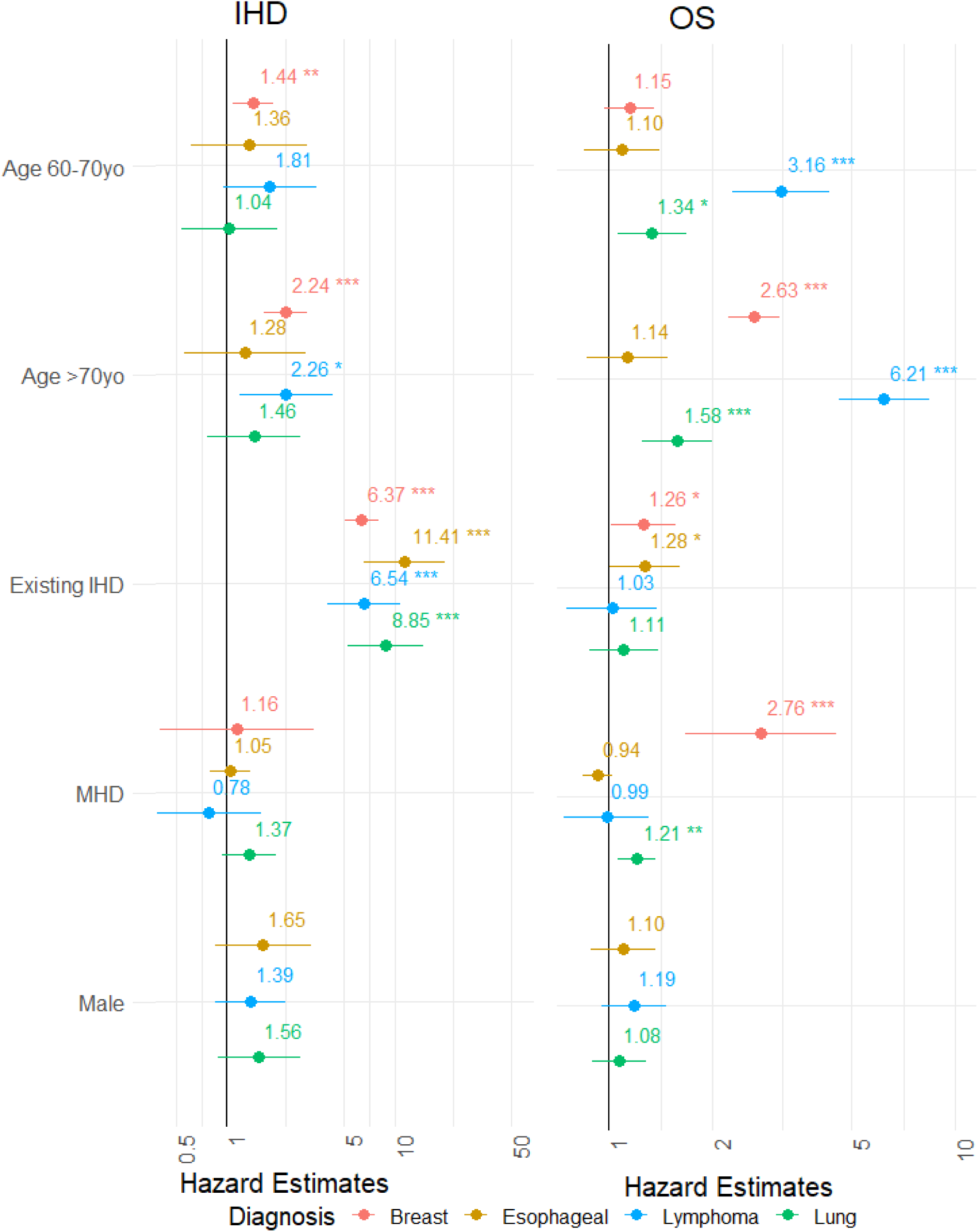
Forest plots of hazard estimates with 95% CI for predictors in eight Cox proportion hazards models on two outcomes - ischemic heart disease (IHD) and overall survival (OS) - separated by four diagnoses. The Breast cancer cohort only includes females. A hazard estimate above one indicates an increased risk of the outcome. Forest plots for overall CVD in supplement Figure A8. P-value thresholds are 0.05(*), 0.01 (**), and 0.001(***).

As sensitivity analyses, we separately analyzed populations with and without existing IHD, but this did not vary greatly from the primary results (Figure A2; Figure A3; Figure A4). Younger patients with breast cancer and lymphoma did show distinct trends (Figure A5; Figure A6). Results for the broader definition of CVD did not differ significantly from IHD (Figure A7; Figure A8). There was substantial correlation observed between dose parameters (Figure A9; Table A3). Adding V5 or V30 to models including MHD therefore did not add significant explainability, and thus results and discussion focus on models without volumetric dose measures.

## Discussion

This study first quantifies mortality and cardiotoxic side effects in thoracically irradiated cancer patients. It then analyzes the relative impact of radiation dose and other patient-specific factors on outcomes. We found the risk of death from cancer far exceeded the risk of cardiac disease, particularly among patients with lung and esophageal cancer. Contrary to earlier findings, no significant relationship was found between cardiac irradiation parameters and subsequent IHD [8–10]. Subsequent IHD risk was dominated by known background population factors of existing IHD, age, and gender in this study. Our findings support that modern RT cardiac sparing advancements limit radiation-related cardiotoxicity and tumor control remains the main clinical problem across all diagnoses studied. Known patient-specific prognostic risk factors impact outcomes significantly more than dose in the achievable cardiac dose range in this series. While these factors are not controllable, they may help identify high-risk patients and inform treatment planning [21].

### Overall Survival

There is a dose-related association between high MHD and poorer OS in breast and lung cancer patients in accordance with other reports in literature [15]. For breast cancer patients, the guidelines define the extent of radiation, particularly parasternal lymph node irradiation, according to disease stage. Therefore, the observed dose relationship with OS may be confounded by patients receiving regional node irradiation having a higher dose to the heart and a less favorable survival of the index cancer. Similarly, different subtypes of lung cancer may tend to occur in different parts of the lungs. Additionally, the size of the primary tumor will influence the risk of irradiating the heart. Therefore, an endpoint specific analysis is necessary for further understanding.

It should surprise no-one that age influences the survival in patients with a favorable cancer prognosis (Figure 4; Figure A5). In addition to the obvious association with general mortality, there may be age-related staging effects and challenges such as poor adherence to treatment protocol. For breast cancer, mammography screening is not recommended after age 70 in Denmark, meaning a higher proportion of patient-detected and thus poorer-prognosis cases [31–33]. For lymphoma, the magnitude of this age effect is much larger than what we observed for breast cancer. The lymphoma data are complex due to confounding with lymphoma subtypes with very different prognoses being diagnosed at different ages and may be treated with radiotherapy at primary treatment or relapse. Nevertheless, the data shows that the prognosis of elderly patients is poor. The systemic treatment for lymphomas, typically developed on younger trial patients, is generally quite aggressive and older patients often do not tolerate this treatment well. The role of radiotherapy is therefore even more important in older patients where the detrimental effects of radiation to the heart need to be carefully balanced against the necessity for adequate treatment of the active cancer.

### Ischemic Heart Disease

Existing IHD was found to increase the risk of subsequent IHD as the strongest predictor in the model (Figure 3). The scale of existing IHD’s impact can be quantified relative to the dose effect. For breast cancer, the presence of existing IHD is equivalent to a MHD increase of 12.5 Gy – doses that should not be seen after contemporary treatment. Similarly, the HR between the oldest age group compared to the youngest is equivalent to an additional 5.4 Gy. Pre-existing disease status and age must therefore be considered when selecting patient interventions, such as late effects surveillance or proton therapy [34,35].

MHD is inversely related to IHD in lymphoma. This result is confounded by the association of age and lymphoma subtype. Younger patients more often have mediastinal disease, typically Hodgkin lymphoma or primary mediastinal large cell lymphoma, which have good prognoses. The combination of young age and good prognoses, even paired with a greater risk of irradiating the heart due to mediastinal RT, seems to confer a lower risk of IHD. However, in a multivariable analysis, this MHD effect on IHD is non-significant.

For breast cancer patients, the observed impact of MHD on OS is unlikely to be mediated by an association with subsequent IHD or CVD. This suggests the dominant effect of OS may be related to the higher disease stage associated with the wider tangential RT fields to cover regional lymph nodes. Prior studies have shown a clear link between MHD and CVD. However, the doses being observed in this contemporary cohort are lower than the doses in previous studies [36]. Guidelines in Denmark were updated to emphasize target coverage over cardiac dose following the 2016 DBCG-IMN’s findings [37]. Our data support this decision. The primary disease should be the main concern and sparing of normal tissue should largely be achieved through technological advances rather than compromising the target dose.

For patients with lung cancer, this study supports the detrimental effect on OS from higher MHD reported by the Manchester group [15]. Yet, we did not observe a corresponding increase in risk of IHD, again putting a question mark over cause and effect. This suggests that survival impacts may be more related to disease characteristics rather than treatment toxicity, as is the case with breast cancer patients. This could include confounding by association between cardiac exposure and mediastinal lymph node involvement or disease extent. Additionally, most lung cancer patients experience recurrence and therefore the competing risk of death due to cancer is high compared to risk of death due to cardiac disease. Access to the specific endpoints of cardiotoxicity improves the interpretability of the dose-response findings compared to assessing OS alone. Our data contributes to quantifying the discussion of the prioritization between target coverage and cardiac risk in lung cancer patients. Still, there is a possibility of significant detrimental effect of radiation to the heart, given the large confidence intervals for the estimate of the effect of dose. Nevertheless, disease control should remain the top priority in lung cancer patients (Figure 5).

### Strengths & Limitations

This study integrates detailed 3D dosimetry on 9,411 patients from a single institution with individually delineated hearts and linked outcome data from Denmark’s national registry systems. The dataset enables the investigation and quantification of cardiotoxic side effects of radiation doses over a 12-year period. Such data can inform the clinically important discussion of balancing exposure of normal tissue against the need to treat the active cancer in patients undergoing radiotherapy with greater confidence than smaller studies with manual data analysis.

A limitation is the study’s lack of chemotherapy data. Cardiotoxicity of some chemotherapy drugs, such as anthracyclines, are well established [38]. Unfortunately, the registries did not allow reliable extraction of chemotherapy data. Similarly, other data, such as diabetes status or BMI, could have been valuable; this data was not reliably available for such a large cohort, but could potentially in the future be extracted using natural language processing of patient charts [39,40]. Additionally, inclusion of staging and tumor characteristics would have added substantial information to a predictive model. Inclusion of coronary artery calcium (CAC) scoring may also contribute to better predictions [41]. Despite these limitations, the model captures important patient related factors combined with dose.

Another limitation is that the median follow-up time of 6.5 years is limited due to data availability within EHR systems. The statistical power is sufficient to solidly resolve conventional risk factors of age and pre-existing disease, but despite 540 events of IHD we do not observe a significant dose response relationship. Supplementary Figure A10 converts our results to excess relative risk (ERR) per Gy for comparison with Darby and Nimwegen and demonstrates overlapping but much wider confidence intervals in this cohort study [9,10]. It may be the narrow range of heart doses in the contemporary patients which challenges the fitting process and limits the power of the current study. Extended follow-up would be advantageous, and the method here could be expanded to larger collaborations for further confirmation.

Future studies could consider target coverage in addition to cardiac dose. For lung cancers, the competing risk of death is so high that a much larger population is needed to detect the effect of MHD on IHD, due to endpoint rarity. The point estimate is still trending as expected, suggesting a trend consistent with observed toxic effects of cardiac exposure [1]. However, the scale of the impact of this aggressive disease significantly overshadows the detrimental impact of radiation on cardiovascular health.

An alternative approach to using whole heart dose metrics is a focus on dose effects on cardiac substructures. Some studies investigating the relationship between cardiac dose, cardiac events and mortality suggest that dose to cardiac substructures is more important than whole heart dose [12,18,42,43]. Zhang et al performed a systematic review of eighteen studies published before 2018 which included locally advanced NSCLC patients treated with concurrent chemo-radiotherapy [44]. All twenty parameters were found to be associated significantly with OS and IHD, the most frequent being MHD, V5, and V30. However, unlike this study, consistent heart dose-volume parameters associated with OS of patients with NSCLC were not identified. Several heart contouring atlases have been published aiming at consistent dose reporting to cardiac substructures [15,16,42,45]. Additionally, multiple open-access AI tools have been published which may allow us to investigate substructure dose trends[46,47]. Furthermore, localization of radio-sensitive regions can be assessed with image-based data mining tools [48–50]. Despite these future prospects, the small magnitude of effect of radiation dose to the whole heart compared to patient related risk factors would suggest the probability of finding a strong association with a substructure of the heart is probably modest.

In conclusion, we have analyzed a large dataset for registry-based assessment of radiation dose effect for the heart. Within the confidence intervals, our dose-effect estimates are consistent with Darby’s famous ERR=7.4 %/Gy, but the risk of cardiac radiation exposure with contemporary treatment techniques is dwarfed by patient-related risk factors and the competing risk of death for most patients. The primary cancer diagnosis remains the single greatest health risk across studied populations, pointing to the importance of reducing cardiac radiation *without* compromising the target coverage.

## Data Availability

Data produced in the present study are available upon reasonable request to the authors.

## Appendix A – Supplementary Tables and Figures

**Table A1.**
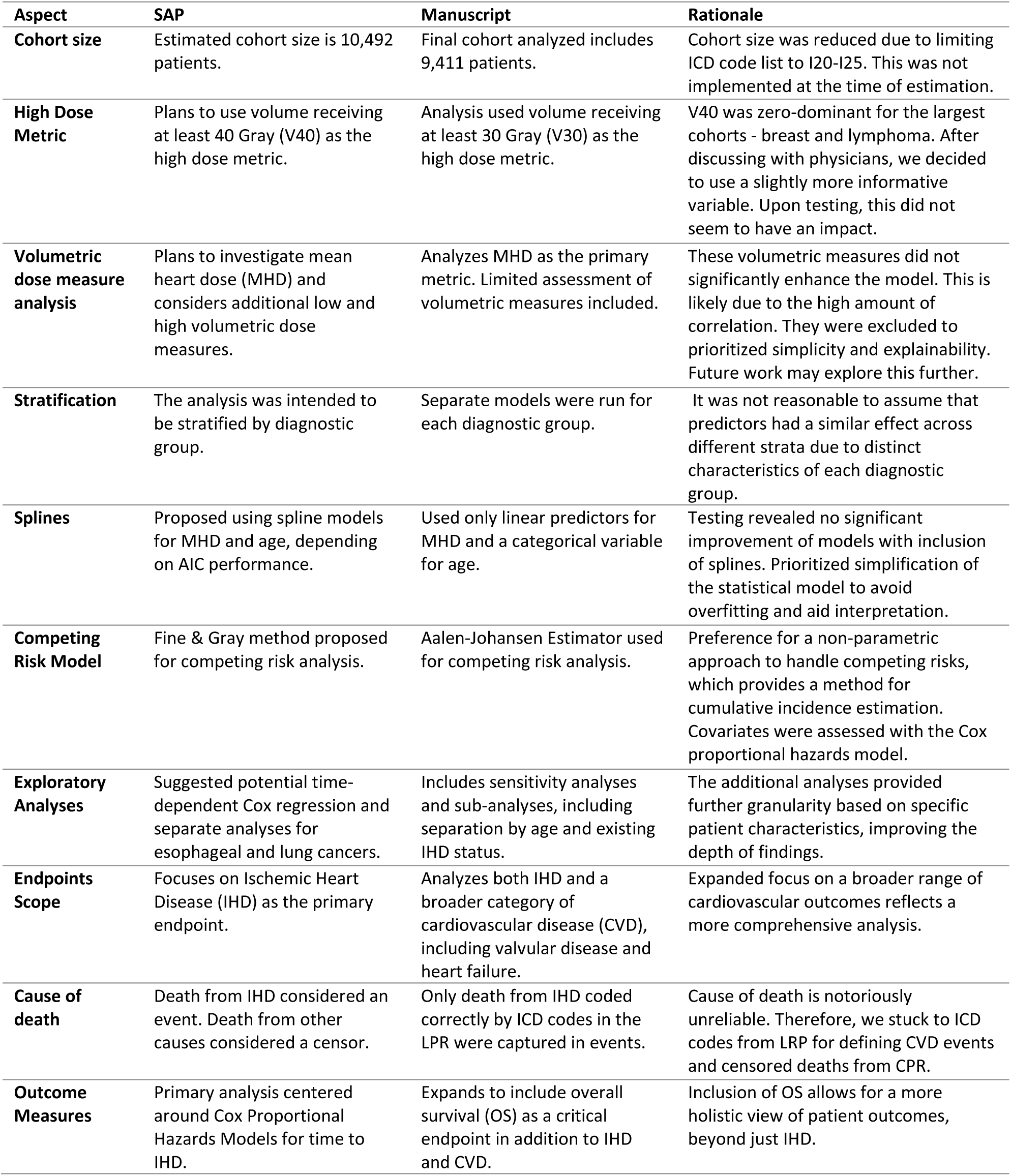
Rationale for changes in methodology from statistical analysis plan (SAP) to manuscript.

| Aspect | SAP | Manuscript | Rationale |
| --- | --- | --- | --- |
| **Cohort size** | Estimated cohort size is 10,492 patients. | Final cohort analyzed includes 9,411 patients. | Cohort size was reduced due to limiting ICD code list to I20-I25. This was not implemented at the time of estimation. |
| **High Dose Metric** | Plans to use volume receiving at least 40 Gray (V40) as the high dose metric. | Analysis used volume receiving at least 30 Gray (V30) as the high dose metric. | V40 was zero-dominant for the largest cohorts - breast and lymphoma. After discussing with physicians, we decided to use a slightly more informative variable. Upon testing, this did not seem to have an impact. |
| **Volumetric dose measure analysis** | Plans to investigate mean heart dose (MHD) and considers additional low and high volumetric dose measures. | Analyzes MHD as the primary metric. Limited assessment of volumetric measures included. | These volumetric measures did not significantly enhance the model. This is likely due to the high amount of correlation. They were excluded to prioritize simplicity and explainability. Future work may explore this further. |
| **Stratification** | The analysis was intended to be stratified by diagnostic group. | Separate models were run for each diagnostic group. | It was not reasonable to assume that predictors had a similar effect across different strata due to distinct characteristics of each diagnostic group. |
| **Splines** | Proposed using spline models for MHD and age, depending on AIC performance. | Used only linear predictors for MHD and a categorical variable for age. | Testing revealed no significant improvement of models with inclusion of splines. Prioritized simplification of the statistical model to avoid overfitting and aid interpretation. |
| **Competing Risk Model** | Fine & Gray method proposed for competing risk analysis. | Aalen-Johansen Estimator used for competing risk analysis. | Preference for a non-parametric approach to handle competing risks, which provides a method for cumulative incidence estimation. Covariates were assessed with the Cox proportional hazards model. |
| **Exploratory Analyses** | Suggested potential time-dependent Cox regression and separate analyses for esophageal and lung cancers. | Includes sensitivity analyses and sub-analyses, including separation by age and existing IHD status. | The additional analyses provided further granularity based on specific patient characteristics, improving the depth of findings. |
| **Endpoints Scope** | Focuses on Ischemic Heart Disease (IHD) as the primary endpoint. | Analyzes both IHD and a broader category of cardiovascular disease (CVD), including valvular disease and heart failure. | Expanded focus on a broader range of cardiovascular outcomes reflects a more comprehensive analysis. |
| **Cause of death** | Death from IHD considered an event. Death from other causes considered a censor. | Only death from IHD coded correctly by ICD codes in the LPR were captured in events. | Cause of death is notoriously unreliable. Therefore, we stuck to ICD codes from LRP for defining CVD events and censored deaths from CPR. |
| **Outcome Measures** | Primary analysis centered around Cox Proportional Hazards Models for time to IHD. | Expands to include overall survival (OS) as a critical endpoint in addition to IHD and CVD. | Inclusion of OS allows for a more holistic view of patient outcomes, beyond just IHD. |

**Table A2.** Number of lymphoma patients with mean heart dose (MHD) > 5 Gy by age group. Chi-squared test for independence yields a p-value < 0.001.

|  | **MHD <5 Gy** | **MHD >5 Gy** |
| --- | --- | --- |
| **<60 years old** | 449 | 103 |
| **60-70 years old** | 285 | 23 |
| **>70 years old** | 359 | 32 |

**Figure A1.**
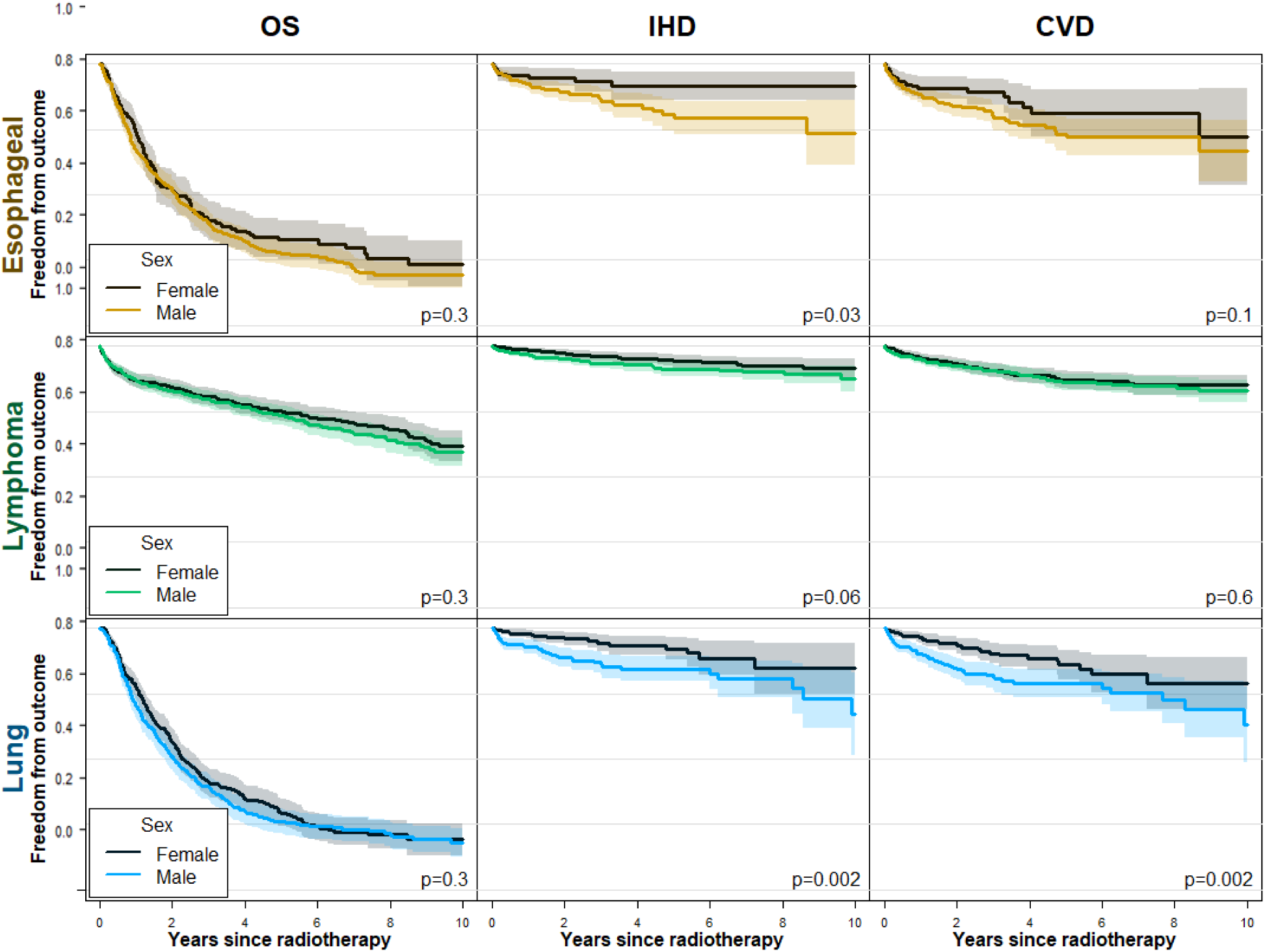
Kaplan-Meier plots for overall survival (OS), ischemic heart disease (IHD), and cardiovascular disease (CVD) by sex in three diagnosis groups. Note: Breast cancer was excluded since only females were analyzed. P-values generated from test for trend.

**Figure A2.**
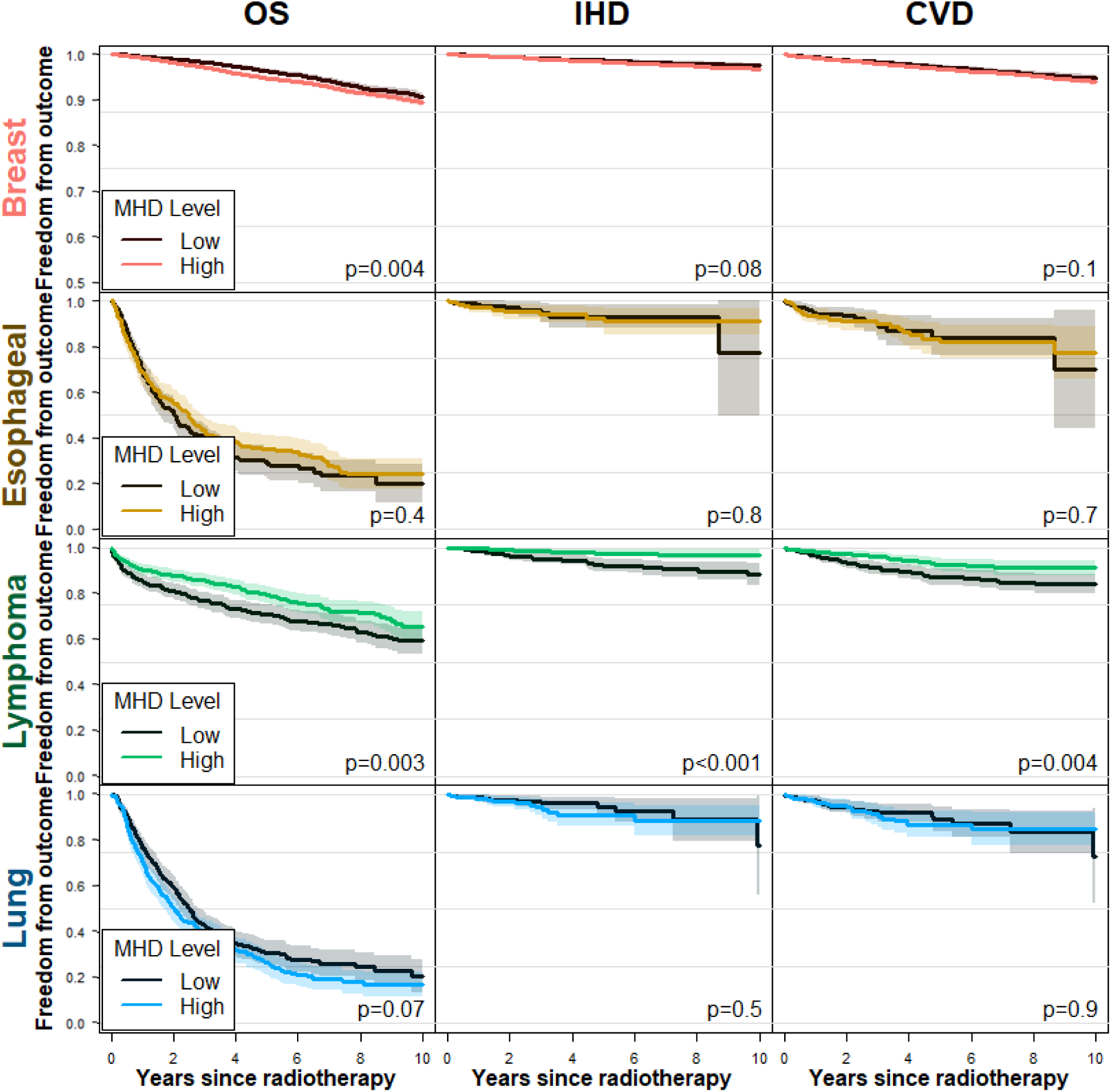
Secondary sub-analyses on populations without existing IHD by diagnosis group. Kaplan-Meier plots for overall survival (OS), ischemic heart disease (IHD), and cardiovascular disease (CVD) by mean heart dose (MHD) level – above and below the group median. P-values generated from test for trend.

**Figure A3.**
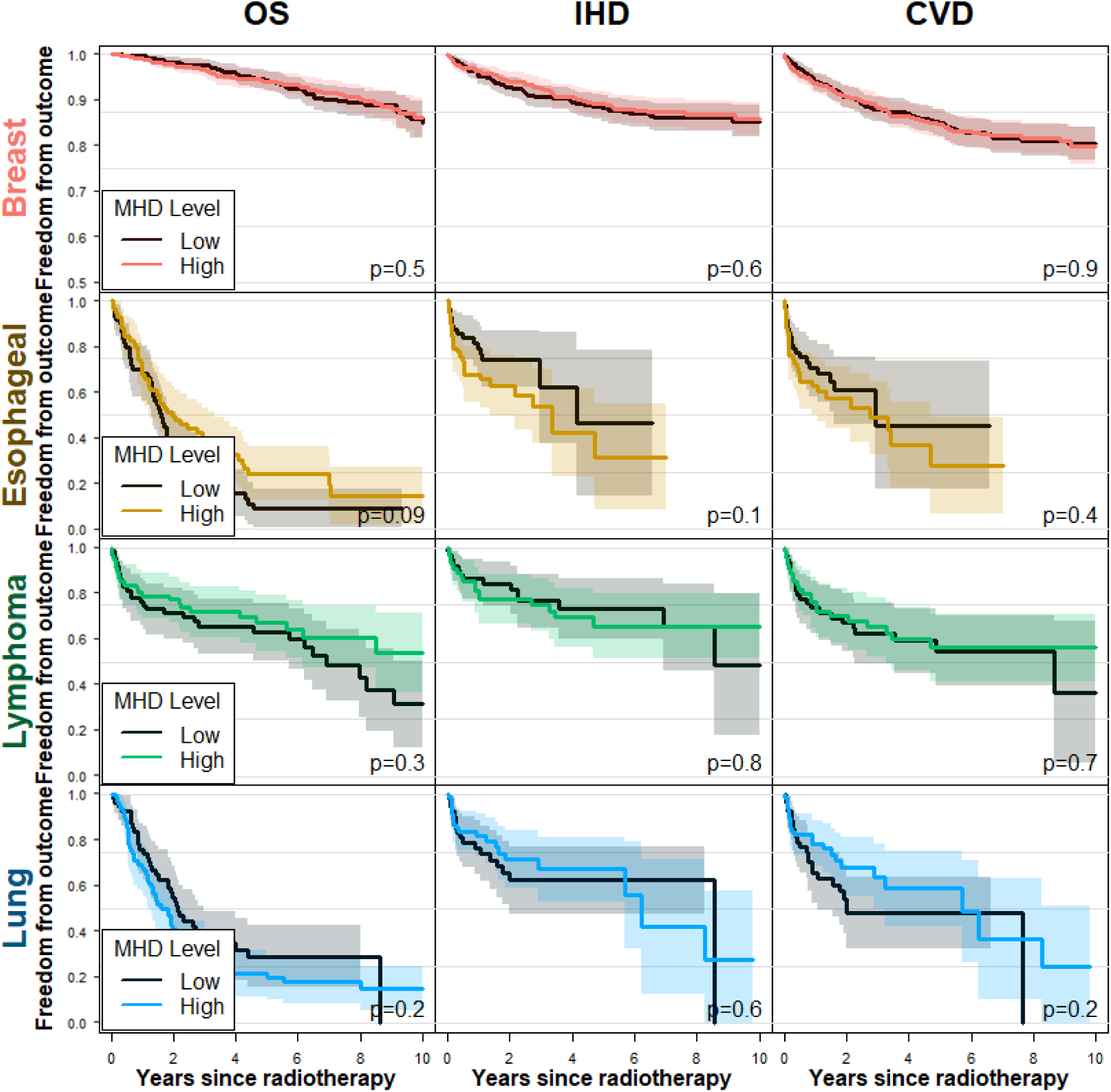
Secondary sub-analyses on populations with existing IHD, by diagnosis group. Kaplan-Meier plots for overall survival (OS), ischemic heart disease (IHD), and cardiovascular disease (CVD) by mean heart dose (MHD) level – above and below the group median. P-values generated from test for trend.

**Figure A4.**
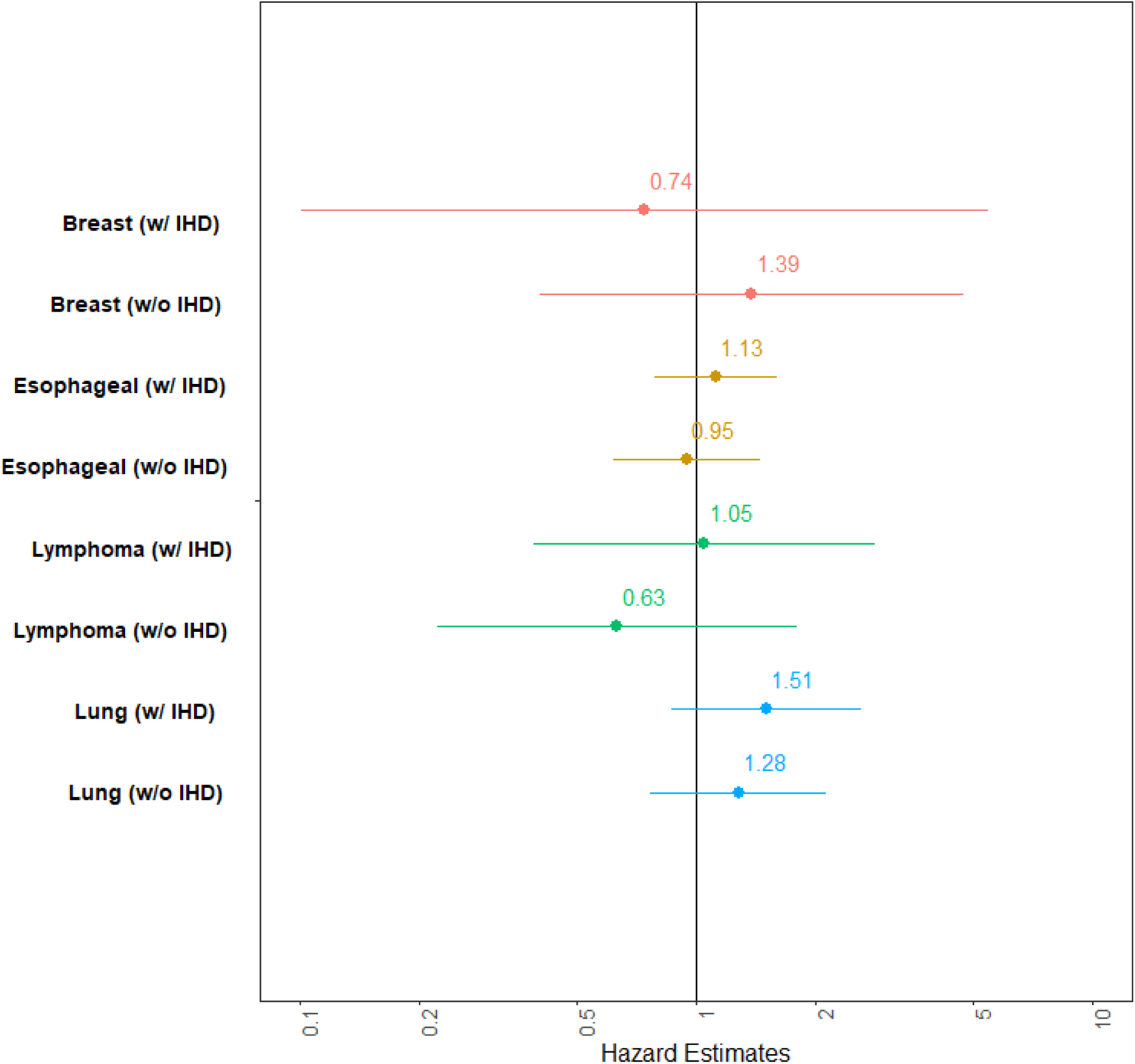
Forest plots hazard estimates with 95% CI for per Gy mean heart dose (MHD) in eight Cox proportion hazards models separated by four diagnoses and existing IHD status (with or without). A hazard estimate above one indicates an increased risk of subsequent IHD.

**Figure A5.**
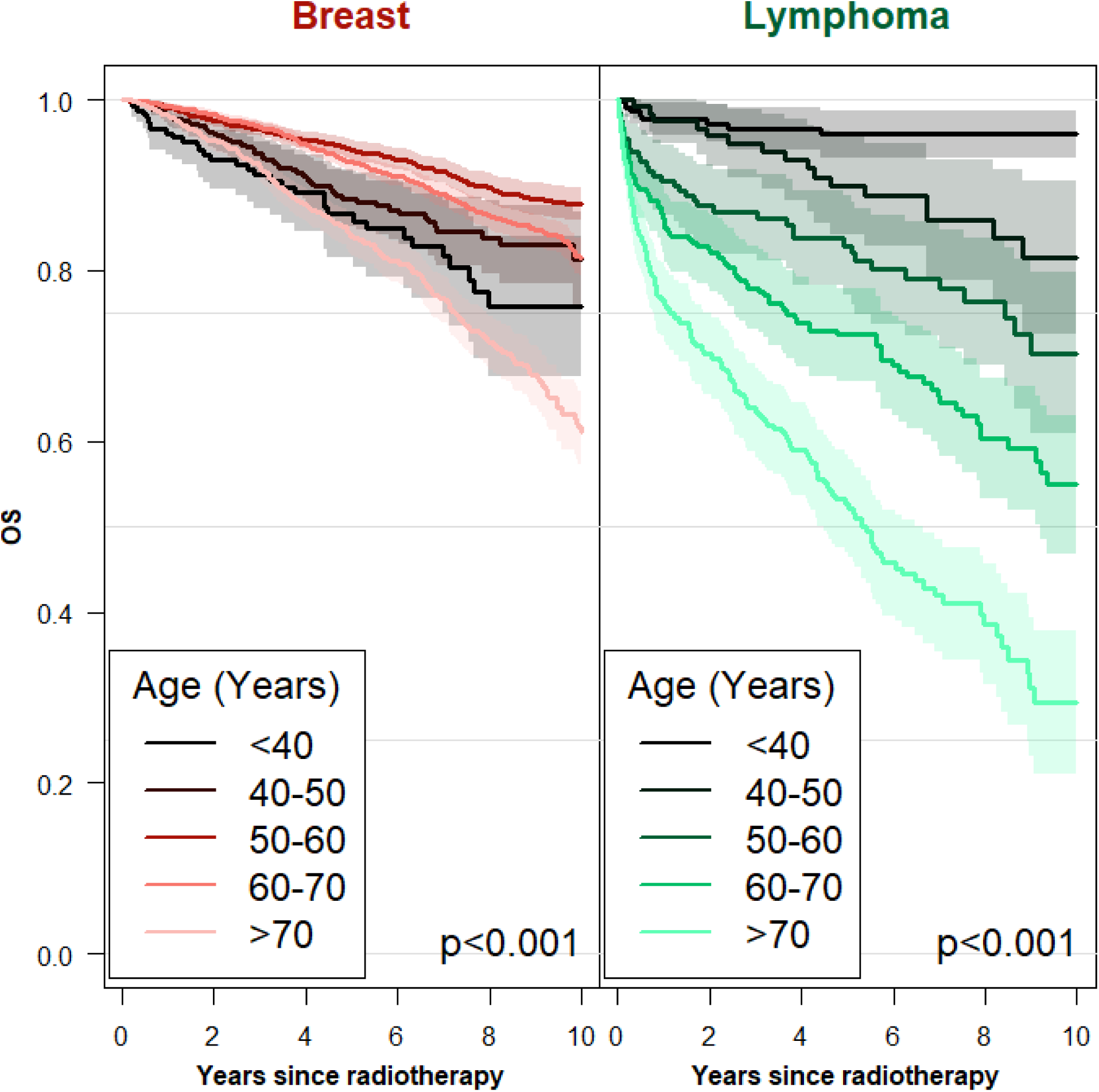
Kaplan-Meier plots for overall survival (OS) by detailed age group (<40, 40-50, 50-60, 60-70, and >70 years old) for breast and lymphoma. P-values generated from test for trend.

**Figure A6.**
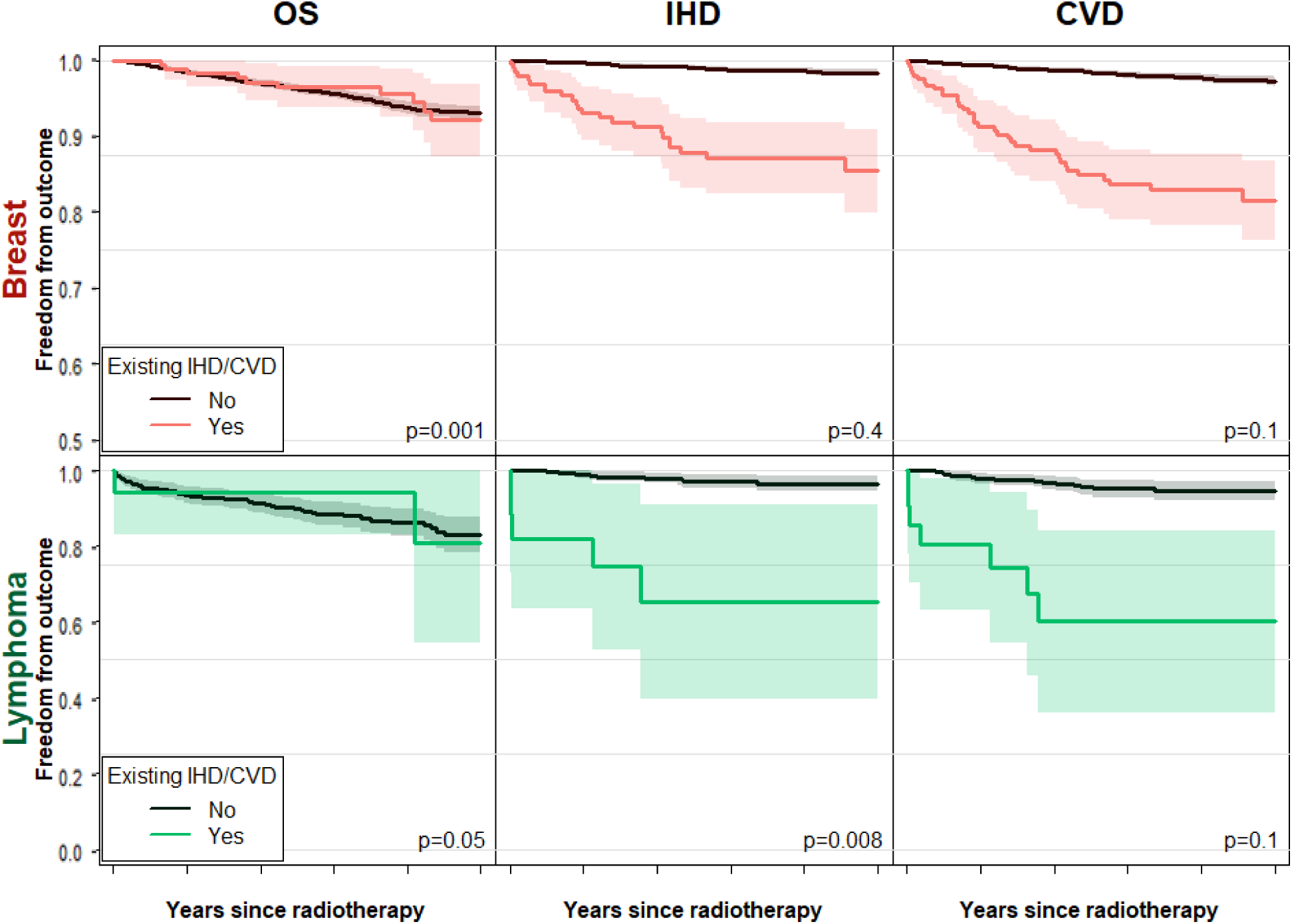
Secondary sub-analysis of youngest patients in the breast and lymphoma cohorts (<60 years old). Kaplan-Meier plots for overall survival (OS) and ischemic heart disease (IHD) by existing IHD as well as cardiovascular disease (CVD) by existing CVD. *Note that the Breast cancer y-axis is truncated at 0.5 to better show separation.* P-values generated from test for trend.

**Figure A7.**
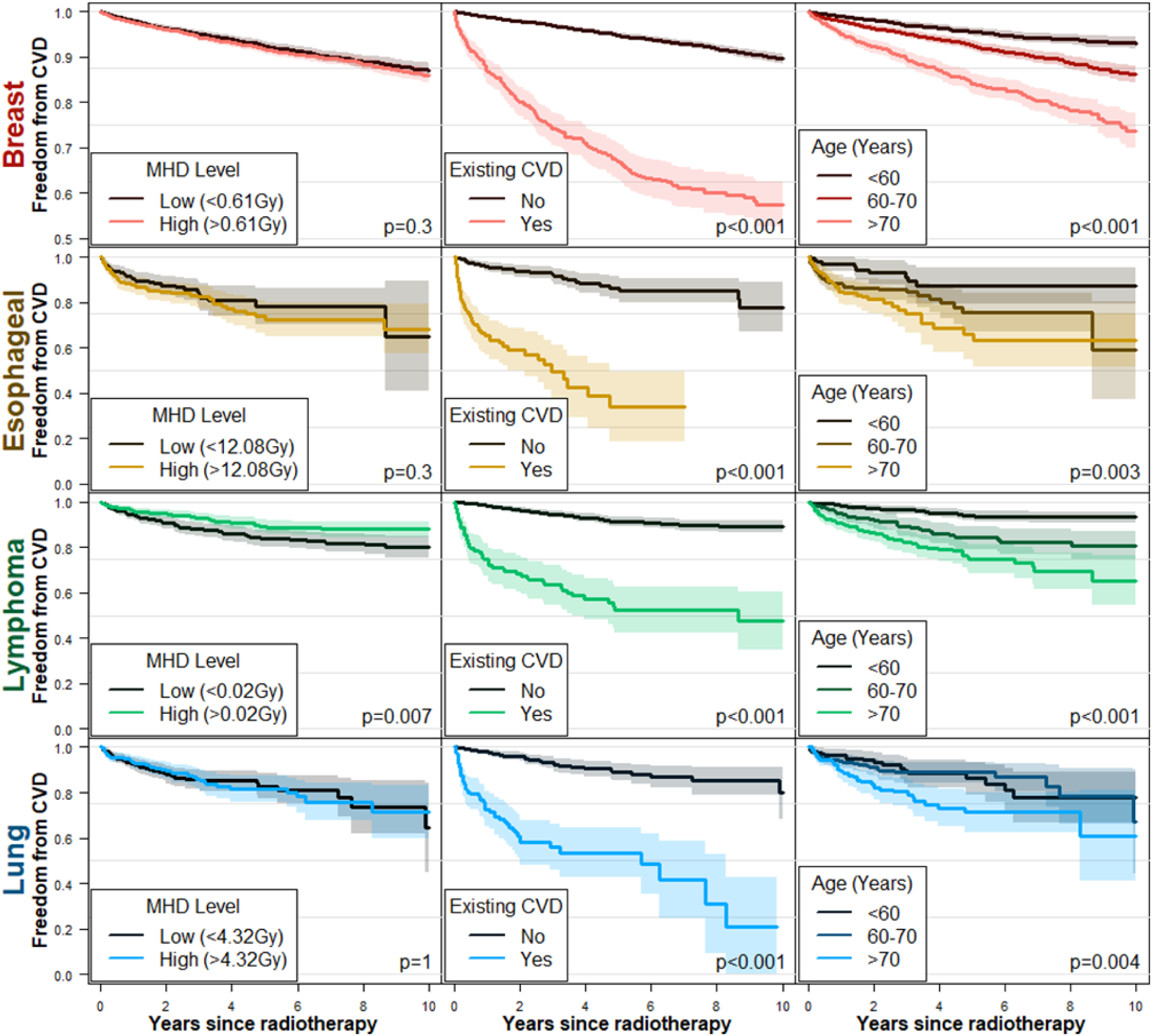
Kaplan-Meier plots for CVD, by mean heart dose (MHD) level (below or above median), existing ischemic heart disease (IHD), and age group (<60, 60-70, and >70 years old), separated by diagnosis. P-values generated from log-rank test (MHD level and existing IHD) and test for trend (age group). Note that the Breast cancer y-axis is truncated at 0.5 to better show separation. Numbers at risk available in supplementary Table A6. For the effect of sex on freedom from CVD see supplementary Figure A1.

**Figure A8.**
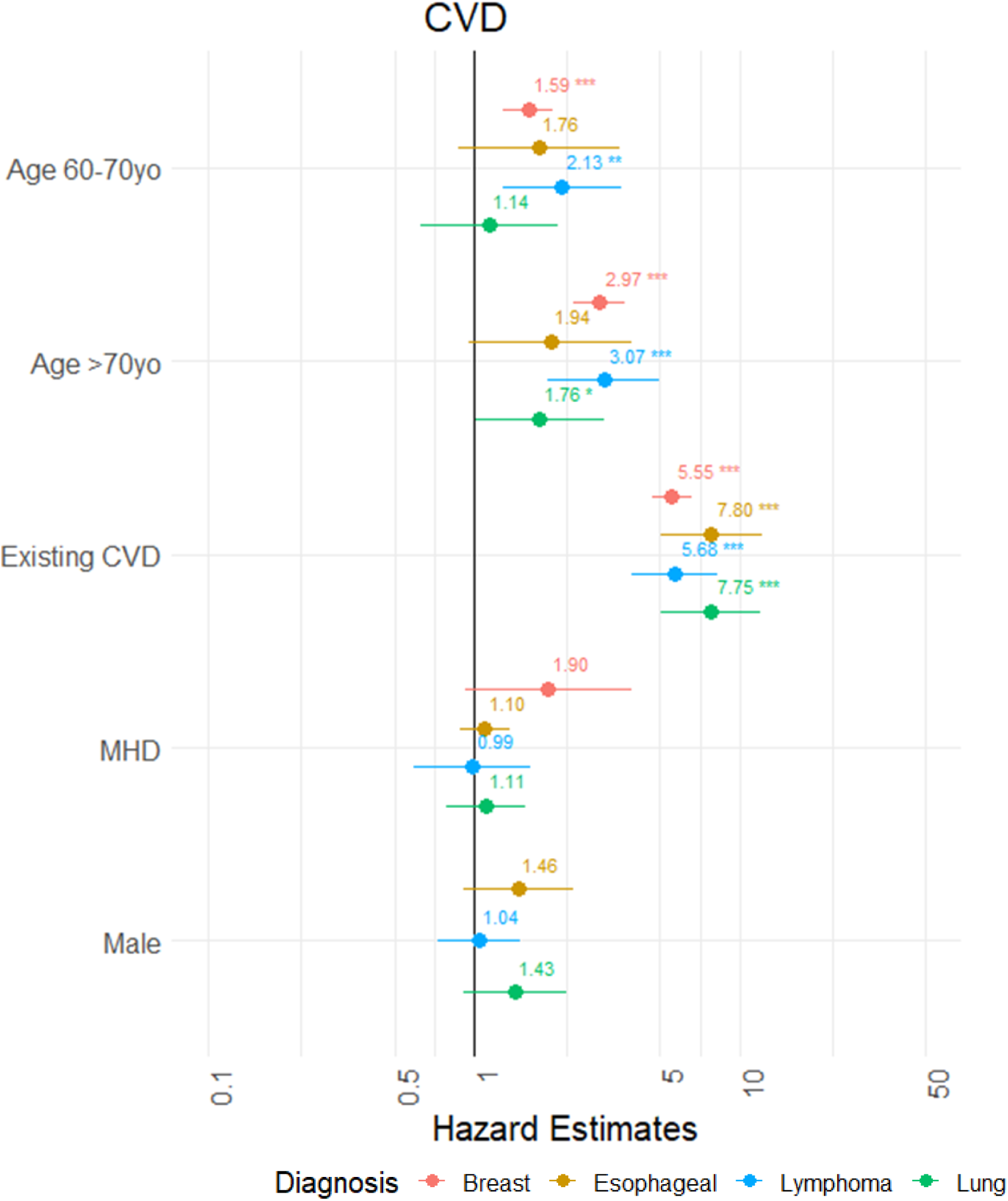
Forest plots of hazard estimates with 95% CI for predictors in Cox proportion hazards models on CVD separated by four diagnoses. The Breast cancer cohort only includes females. A hazard estimate above one indicates an increased risk of the outcome. P-value thresholds are 0.05(*), 0.01 (**), and 0.001(***).

**Table A3.** Kendall correlations between mean heart dose (MHD) and low (V5) and high (V30) volumetric dose measures, by diagnosis group.

|  | MHD X V5 | MHD X V30 | V5 X V30 |
| --- | --- | --- | --- |
| **Breast** | 0.775 | 0.636 | 0.745 |
| **Esophageal** | 0.684 | 0.833 | 0.661 |
| **Lung** | 0.812 | 0.808 | 0.703 |
| **Lymphoma** | 0.727 | 0.581 | 0.748 |

**Figure A9.**
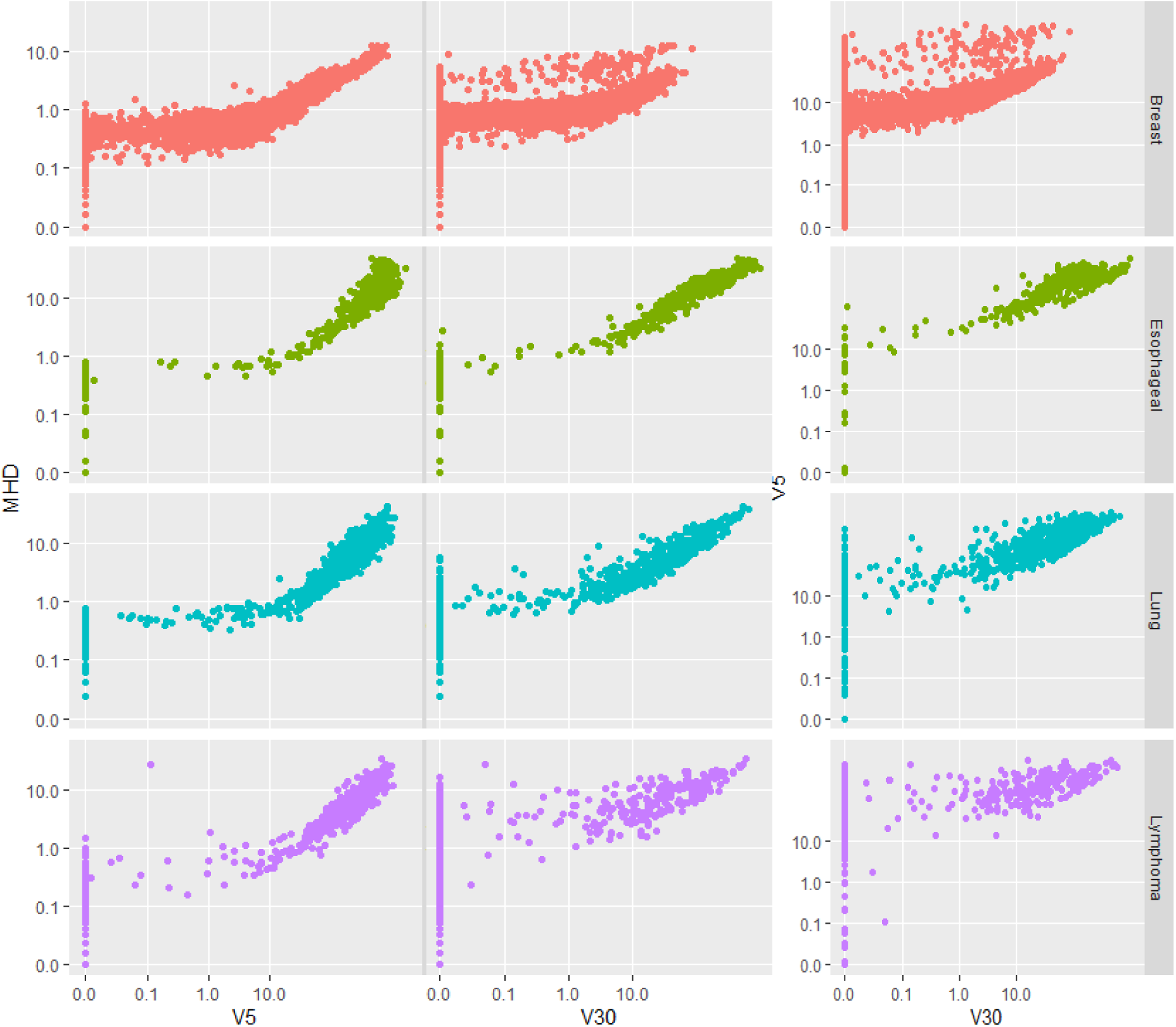
Scatter plot of mean heart dose (MHD) and low (V5) and high (V30) volumetric dose measures, by diagnosis group

**Table A4.**
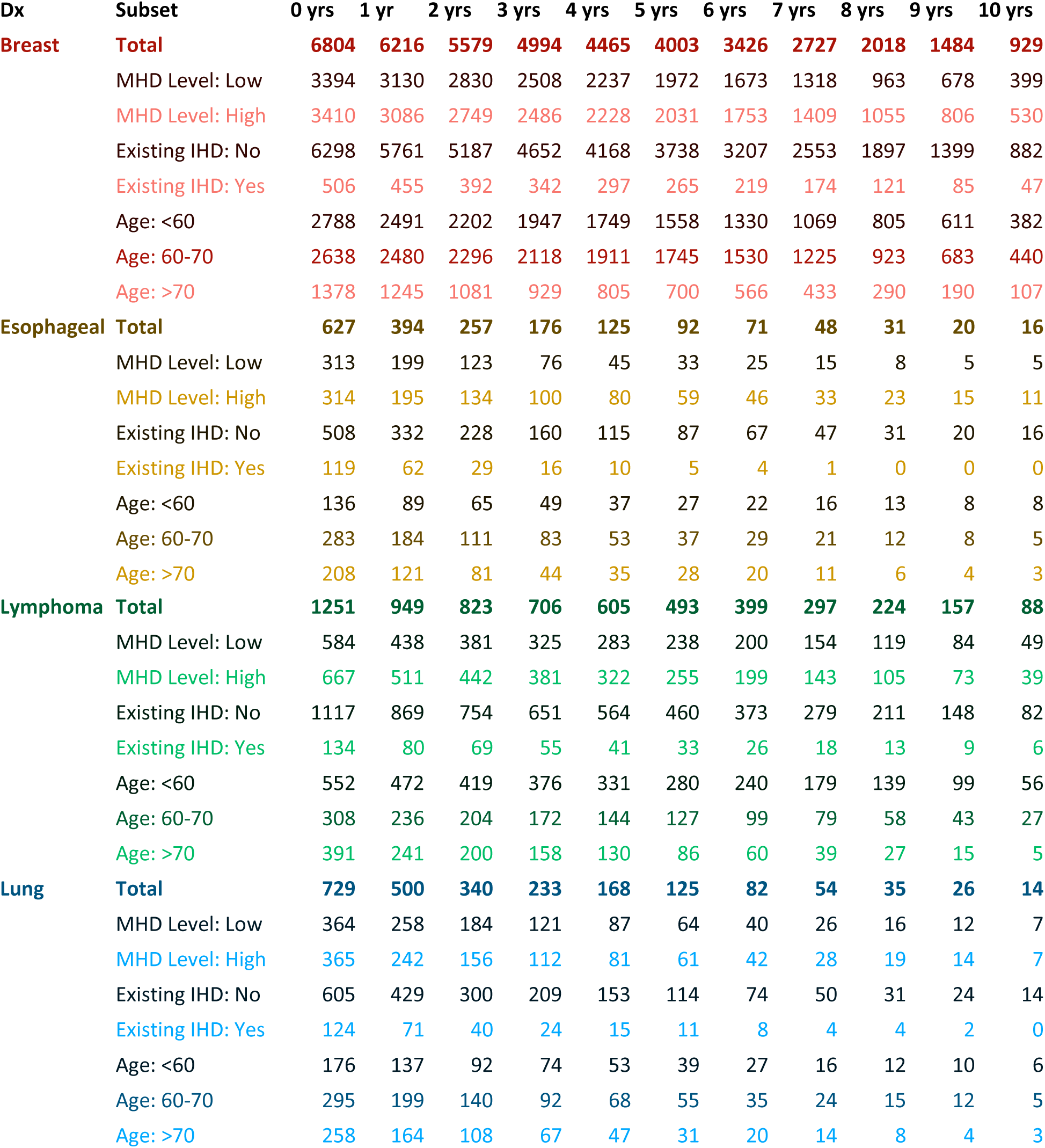
Numbers at risk for ischemic heart disease (IHD) shown in Figure 3.

| Dx | Subset | 0 yrs | 1 yr | 2 yrs | 3 yrs | 4 yrs | 5 yrs | 6 yrs | 7 yrs | 8 yrs | 9 yrs | 10 yrs |
| --- | --- | --- | --- | --- | --- | --- | --- | --- | --- | --- | --- | --- |
| **Breast** | **Total** | **6804** | **6216** | **5579** | **4994** | **4465** | **4003** | **3426** | **2727** | **2018** | **1484** | **929** |
|  | MHD Level: Low | 3394 | 3130 | 2830 | 2508 | 2237 | 1972 | 1673 | 1318 | 963 | 678 | 399 |
|  | MHD Level: High | 3410 | 3086 | 2749 | 2486 | 2228 | 2031 | 1753 | 1409 | 1055 | 806 | 530 |
|  | Existing IHD: No | 6298 | 5761 | 5187 | 4652 | 4168 | 3738 | 3207 | 2553 | 1897 | 1399 | 882 |
|  | Existing IHD: Yes | 506 | 455 | 392 | 342 | 297 | 265 | 219 | 174 | 121 | 85 | 47 |
|  | Age: <60 | 2788 | 2491 | 2202 | 1947 | 1749 | 1558 | 1330 | 1069 | 805 | 611 | 382 |
|  | Age: 60-70 | 2638 | 2480 | 2296 | 2118 | 1911 | 1745 | 1530 | 1225 | 923 | 683 | 440 |
|  | Age: >70 | 1378 | 1245 | 1081 | 929 | 805 | 700 | 566 | 433 | 290 | 190 | 107 |
| **Esophageal** | **Total** | **627** | **394** | **257** | **176** | **125** | **92** | **71** | **48** | **31** | **20** | **16** |
|  | MHD Level: Low | 313 | 199 | 123 | 76 | 45 | 33 | 25 | 15 | 8 | 5 | 5 |
|  | MHD Level: High | 314 | 195 | 134 | 100 | 80 | 59 | 46 | 33 | 23 | 15 | 11 |
|  | Existing IHD: No | 508 | 332 | 228 | 160 | 115 | 87 | 67 | 47 | 31 | 20 | 16 |
|  | Existing IHD: Yes | 119 | 62 | 29 | 16 | 10 | 5 | 4 | 1 | 0 | 0 | 0 |
|  | Age: <60 | 136 | 89 | 65 | 49 | 37 | 27 | 22 | 16 | 13 | 8 | 8 |
|  | Age: 60-70 | 283 | 184 | 111 | 83 | 53 | 37 | 29 | 21 | 12 | 8 | 5 |
|  | Age: >70 | 208 | 121 | 81 | 44 | 35 | 28 | 20 | 11 | 6 | 4 | 3 |
| **Lymphoma** | **Total** | **1251** | **949** | **823** | **706** | **605** | **493** | **399** | **297** | **224** | **157** | **88** |
|  | MHD Level: Low | 584 | 438 | 381 | 325 | 283 | 238 | 200 | 154 | 119 | 84 | 49 |
|  | MHD Level: High | 667 | 511 | 442 | 381 | 322 | 255 | 199 | 143 | 105 | 73 | 39 |
|  | Existing IHD: No | 1117 | 869 | 754 | 651 | 564 | 460 | 373 | 279 | 211 | 148 | 82 |
|  | Existing IHD: Yes | 134 | 80 | 69 | 55 | 41 | 33 | 26 | 18 | 13 | 9 | 6 |
|  | Age: <60 | 552 | 472 | 419 | 376 | 331 | 280 | 240 | 179 | 139 | 99 | 56 |
|  | Age: 60-70 | 308 | 236 | 204 | 172 | 144 | 127 | 99 | 79 | 58 | 43 | 27 |
|  | Age: >70 | 391 | 241 | 200 | 158 | 130 | 86 | 60 | 39 | 27 | 15 | 5 |
| **Lung** | **Total** | **729** | **500** | **340** | **233** | **168** | **125** | **82** | **54** | **35** | **26** | **14** |
|  | MHD Level: Low | 364 | 258 | 184 | 121 | 87 | 64 | 40 | 26 | 16 | 12 | 7 |
|  | MHD Level: High | 365 | 242 | 156 | 112 | 81 | 61 | 42 | 28 | 19 | 14 | 7 |
|  | Existing IHD: No | 605 | 429 | 300 | 209 | 153 | 114 | 74 | 50 | 31 | 24 | 14 |
|  | Existing IHD: Yes | 124 | 71 | 40 | 24 | 15 | 11 | 8 | 4 | 4 | 2 | 0 |
|  | Age: <60 | 176 | 137 | 92 | 74 | 53 | 39 | 27 | 16 | 12 | 10 | 6 |
|  | Age: 60-70 | 295 | 199 | 140 | 92 | 68 | 55 | 35 | 24 | 15 | 12 | 5 |
|  | Age: >70 | 258 | 164 | 108 | 67 | 47 | 31 | 20 | 14 | 8 | 4 | 3 |

**Table A5.**
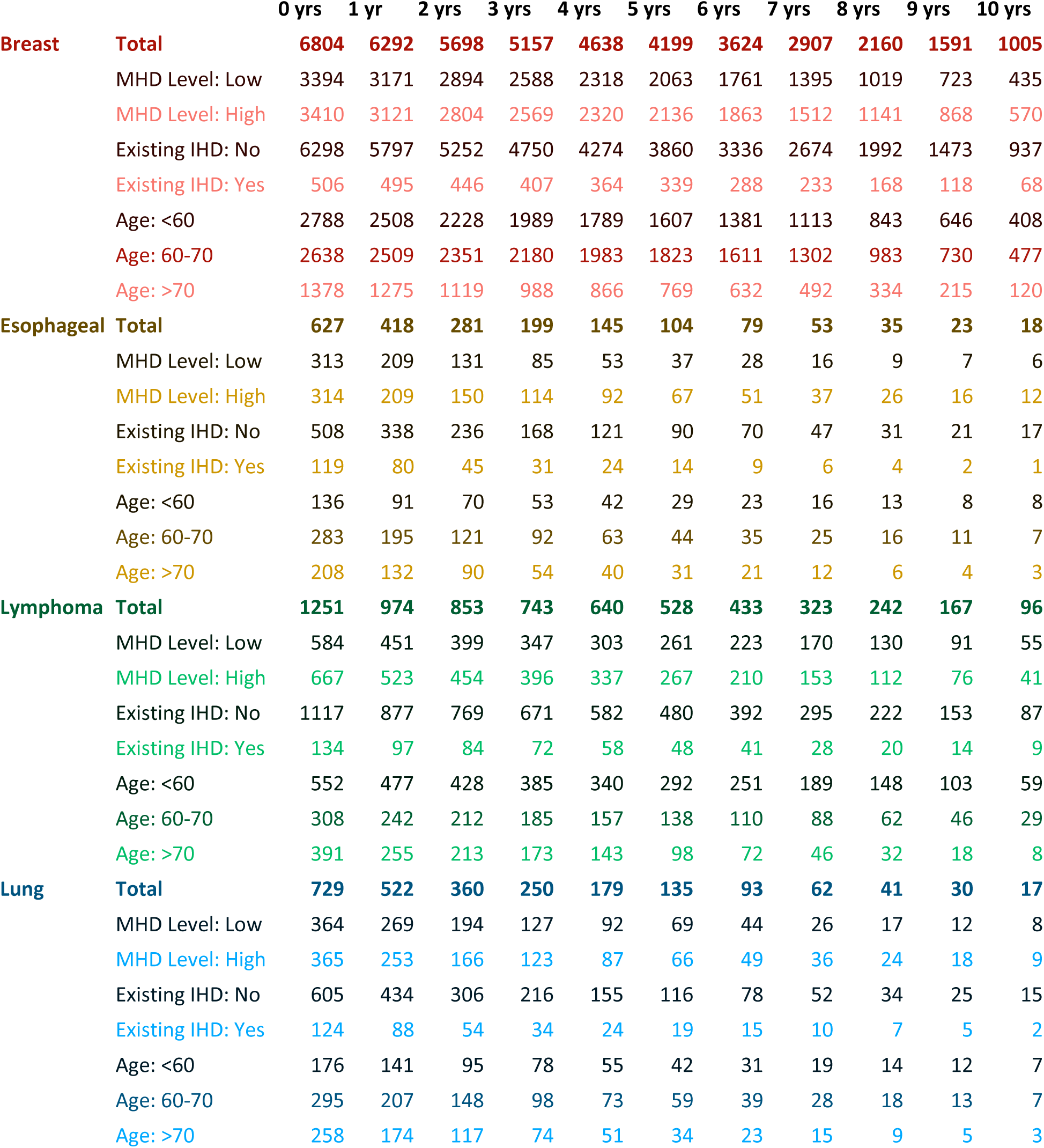
Numbers at risk for overall survival (OS) shown in Figure 4.

**Table A6.** Numbers at risk for cardiovascular disease (CVD) shown in Figure A7.

|  |  | 0 yrs | 1 yr | 2 yrs | 3 yrs | 4 yrs | 5 yrs | 6 yrs | 7 yrs | 8 yrs | 9 yrs | 10 yrs |
| --- | --- | --- | --- | --- | --- | --- | --- | --- | --- | --- | --- | --- |
| **Breast** | **Total** | **6804** | **6216** | **5579** | **4994** | **4465** | **4003** | **3426** | **2727** | **2018** | **1484** | **929** |
|  | MHD Level: Low | 3394 | 3103 | 2789 | 2464 | 2190 | 1916 | 1627 | 1275 | 926 | 653 | 385 |
|  | MHD Level: High | 3410 | 3044 | 2697 | 2434 | 2175 | 1968 | 1694 | 1363 | 1014 | 775 | 511 |
|  | Existing CVD: No | 6186 | 5621 | 5045 | 4523 | 4043 | 3601 | 3091 | 2455 | 1813 | 1339 | 849 |
|  | Existing CVD: Yes | 618 | 526 | 441 | 375 | 322 | 283 | 230 | 183 | 127 | 89 | 47 |
|  | Age: <60 | 2788 | 2479 | 2183 | 1926 | 1726 | 1533 | 1306 | 1046 | 787 | 599 | 376 |
|  | Age: 60-70 | 2638 | 2457 | 2262 | 2081 | 1871 | 1696 | 1479 | 1182 | 884 | 652 | 420 |
|  | Age: >70 | 1378 | 1211 | 1041 | 891 | 768 | 655 | 536 | 410 | 269 | 177 | 100 |
| **Esophageal** | **Total** | **627** | **394** | **257** | **176** | **125** | **92** | **71** | **48** | **31** | **20** | **16** |
|  | MHD Level: Low | 313 | 190 | 117 | 71 | 41 | 30 | 23 | 14 | 8 | 5 | 5 |
|  | MHD Level: High | 314 | 186 | 129 | 96 | 74 | 56 | 44 | 33 | 23 | 14 | 11 |
|  | Existing CVD: No | 491 | 312 | 214 | 150 | 104 | 80 | 63 | 46 | 31 | 19 | 16 |
|  | Existing CVD: Yes | 136 | 64 | 32 | 17 | 11 | 6 | 4 | 1 | 0 | 0 | 0 |
|  | Age: <60 | 136 | 88 | 65 | 49 | 37 | 27 | 22 | 16 | 13 | 8 | 8 |
|  | Age: 60-70 | 283 | 176 | 104 | 76 | 48 | 35 | 27 | 20 | 12 | 7 | 5 |
|  | Age: >70 | 208 | 112 | 77 | 42 | 30 | 24 | 18 | 11 | 6 | 4 | 3 |
| **Lymphoma** | **Total** | **1251** | **949** | **823** | **706** | **605** | **493** | **399** | **297** | **224** | **157** | **88** |
|  | MHD Level: Low | 584 | 427 | 365 | 309 | 268 | 227 | 193 | 149 | 114 | 80 | 48 |
|  | MHD Level: High | 667 | 504 | 433 | 371 | 308 | 241 | 188 | 138 | 101 | 72 | 39 |
|  | Existing CVD: No | 1085 | 843 | 727 | 623 | 534 | 437 | 357 | 268 | 202 | 142 | 80 |
|  | Existing CVD: Yes | 166 | 88 | 71 | 57 | 42 | 31 | 24 | 19 | 13 | 10 | 7 |
|  | Age: <60 | 552 | 469 | 414 | 371 | 324 | 273 | 235 | 176 | 136 | 98 | 55 |
|  | Age: 60-70 | 308 | 230 | 197 | 164 | 133 | 117 | 91 | 75 | 55 | 42 | 27 |
|  | Age: >70 | 391 | 232 | 187 | 145 | 119 | 78 | 55 | 36 | 24 | 12 | 5 |
| **Lung** | **Total** | **729** | **500** | **340** | **233** | **168** | **125** | **82** | **54** | **35** | **26** | **14** |
|  | MHD Level: Low | 364 | 251 | 176 | 115 | 85 | 62 | 40 | 26 | 15 | 12 | 7 |
|  | MHD Level: High | 365 | 240 | 153 | 112 | 79 | 59 | 41 | 27 | 19 | 14 | 7 |
|  | Existing CVD: No | 590 | 418 | 289 | 202 | 149 | 110 | 72 | 49 | 31 | 24 | 14 |
|  | Existing CVD: Yes | 139 | 73 | 40 | 25 | 15 | 11 | 9 | 4 | 3 | 2 | 0 |
|  | Age: <60 | 176 | 136 | 89 | 72 | 53 | 39 | 27 | 16 | 12 | 10 | 6 |
|  | Age: 60-70 | 295 | 195 | 137 | 90 | 67 | 54 | 35 | 24 | 14 | 12 | 5 |
|  | Age: >70 | 258 | 160 | 103 | 65 | 44 | 28 | 19 | 13 | 8 | 4 | 3 |

**Figure A10.**
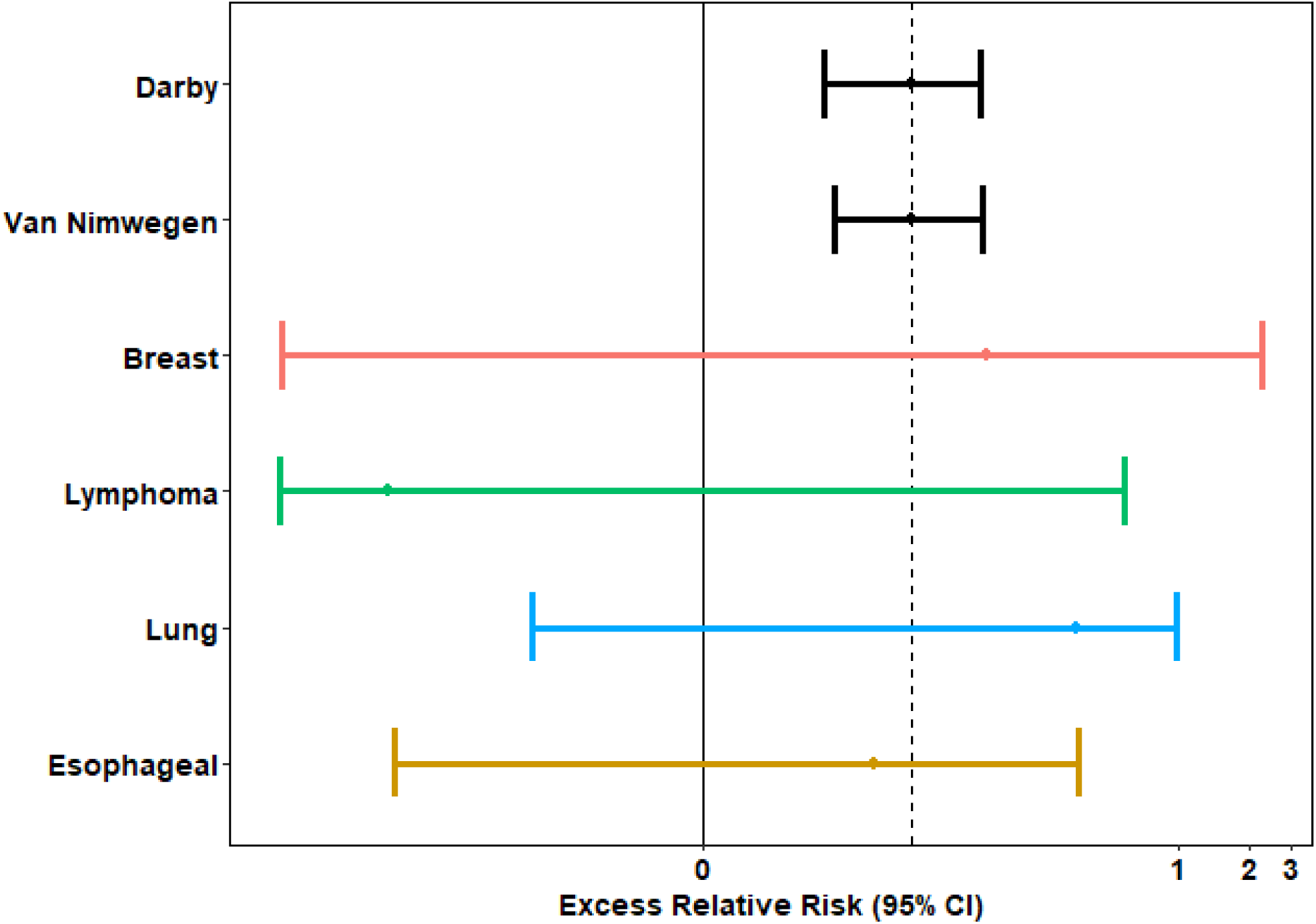
Converted excess relative risk (and 95% confidence intervals) of per Gy mean heart dose by diagnosis and compared with existing literature (Darby and Van Nimwegen) [9,10]. The x-axis is displayed on a pseudo logarithmic scale.

